# Identifying shared and parasite-specific determinants of *Plasmodium falciparum* and *Schistosoma mansoni* co-infection in a large-scale community-based study in rural Uganda

**DOI:** 10.64898/2026.10.29.26366722

**Authors:** Max M. Lang, Violet Tuhaise, Emily Asiimwe, Ronald Bogere, Asmin Mohamed, Babi Isma, Jovan D. Batte, Betty Nabatte, Narcis B. Kabatereine, Christl A. Donnelly, Goylette F. Chami

## Abstract

Malaria and schistosomiasis are co-endemic across much of sub-Saharan Africa, yet it remains unclear whether co-infections reflect shared risk factors, unexplained biological determinants, or chance co-occurrence. We analysed data collected at recruitment from 4290 individuals aged 5–90 years across 52 villages in rural Uganda within the SchistoTrack cohort. A bivariate copula model was developed to separate shared and disease-specific associations from residual dependence. Prevalence was 41.47% for *Schistosoma mansoni*, 41.19% for *Plasmodium falciparum*, and 18.51% for co-infection. Prevalence overlapped in school-aged children with co-infection prevalence peaking at 13 years. Greater distance to a government health centre was the strongest shared determinant, apart from age, male gender and living in western districts, increasing the risk of both. Local water ecology influenced each pathogen in opposing directions. Fishing was associated with higher *S. mansoni* and co-infection risks, but not *P. falciparum* risk. A modelled scenario combining household drinking-water treatment, handwashing facilities and protected water sources with cessation of freshwater contact reduced co-infection prevalence by 32%. Our findings support integrated control around shared demographics and limited healthcare access and household water infrastructure, while highlighting the need for non-integrated approaches for ecological interventions aiming to reduce parasite-specific transmission determinants.

## Introduction

Malaria and schistosomiasis are two of the most common parasitic diseases across sub-Saharan Africa with an estimated 500 million people living in co-endemic areas [1, 2, 3]. Co-infection, defined here as concurrent infection with both parasites in the same individual, has been linked to anaemia and impaired growth in children [4, 5]. Individuals living in co-endemic areas often have limited access to health services and to water, sanitation and hygiene (WASH) infrastructure [5]. Integration of schistosomiasis control with malaria programmes has been advocated since at least 2005 [6, 7], and the latest World Health Organization (WHO) neglected tropical disease (NTD) roadmap 2030 emphasises the need for integrated control [8, 2]. Integration can proceed by targeting shared determinants, so that co-infection falls as both infections decline, or by identifying determinants of co-occurrence itself, so that programmes reach individuals carrying both parasites directly. Which route is more appropriate depends on whether substantial dependence between the two infections remains after accounting for measured shared and disease-specific determinants. Because both infections are endemic in sub-Saharan Africa, a fraction of co-infection is expected by chance alone [4, 9, 5]. However, additional co-infections can arise if the same individuals share risk factors such as treatment being unavailable or similar demographics, or if there is biological dependence where one infection might alter susceptibility to the other [10, 11]. Yet, for malaria and schistosomiasis, which determinants are shared and which are disease-specific has not been established [12]. Further, how much dependence remains once such determinants are accounted for is unknown, despite being essential for effective integrated control [13, 4]. This study focuses on identifying parasite-specific and shared risk factors for *Plasmodium falciparum* and *Schistosoma mansoni* infection.

Although malaria and schistosomiasis overlap geographically, their transmission routes fundamentally differ. *P. falciparum* is transmitted by a mosquito vector, whereas *S. mansoni* is acquired through contact with freshwater harbouring cercariae shed by intermediate-host snails [14, 15, 3, 16]. For example, in lakeside communities of rural Uganda, malaria is caused predominantly by *P. falciparum* [17], transmitted by *Anopheles* mosquitoes that breed in vegetated standing water near dwellings, whereas intestinal schistosomiasis is caused by *S. mansoni*, whose intermediate hosts are freshwater snails of the genus *Biomphalaria* [18]. Both species lead to severe morbidity in this region. *P. falciparum* is the main cause of severe malaria and anaemia [19], while chronic *S. mansoni* infection leads to periportal fibrosis and portal hypertension [20].

Existing malaria–schistosome co-infection investigations have focused on limited populations, mostly drawn from schools and health clinics, with nearly no quantification of shared risk factors and community-level risk profiles unexplored [12]. Co-infection risk is highest in older children and adolescents around 8–17 years of age [21, 11, 4], and pooled estimates suggest higher odds among males in community settings [12]. Yet, treatment campaigns for malaria and schistosomiasis have much narrower age targets. Malaria control has historically concentrated on children under five years of age [19] and in Uganda community-based integrated case management is still restricted to this age group, although the national strategy plans to extend it to all ages [17, 22]. Preventive chemotherapy for schistosomiasis, by contrast, is delivered mainly to school-aged children between roughly 5 and 14 years, typically through school-based mass drug administration (MDA) [23].

Each parasite has a distinct ecological niche [15, 18], so conditions that decrease transmission of one parasite may sustain or increase transmission of the other, with implications for the effectiveness of any shared environmental intervention [16]. Prevention therefore depends on identifying which determinants are shared and which act in opposite directions. Beyond prevention, integrated control also concerns case management. Integrated treatment for co-infection may be agnostic of shared risk factors if diagnostics are readily available to screen all individuals and there is no need for targeting who is diagnosed [19, 24]. In the absence of blanket diagnostics, demographic profiles would be needed at the least to target likely co-infected individuals. Case management may also need to change if *S. mansoni* alters *P. falciparum* parasitaemia [10, 11], or if co-infected individuals carry a greater burden of anaemia [24, 25]. Each of these hypothetical scenarios remains unaddressed without separating shared from parasite-specific determinants.

Co-infection has thus far been studied only as a binary outcome, ignoring the representation of the two pathogens as two dependent outcomes. As such, empirical evidence on the joint distribution of the two infections is scarce [9, 12]. Pooled estimates nonetheless show co-infection occurring more often than chance, though with substantial heterogeneity. Schistosome-infected individuals were around 27% more likely to also be infected with malaria [12]. Methodologically, this crude excess cannot be interpreted without separating shared determinants from residual dependence, and existing co-infection analyses with binary outcomes cannot make that separation. All 30 studies in the recent review collapsed four possible joint infection states (uninfected, malaria only, schistosomiasis only, co-infected) into a single binary outcome and primarily fitted univariate logistic regression [12]. A broader review of multi-response methods for co-infection found that most epidemiological studies use multinomial models, which similarly conflate marginal effects with cross-pathogen dependence [13]. Neither univariate logistic nor multinomial regression can identify which determinants are shared between pathogens, which determinants are pathogen-specific, or whether residual dependence remains after accounting for shared determinants.

We examined 4290 individuals from the SchistoTrack cohort [26, 27], a population-based prospective cohort study spanning 52 villages across three rural districts of Uganda. Cohort participants were aged 5 to 90 years. Malaria infection was ascertained by rapid diagnostic test (RDT), with almost all infections being *P. falciparum* [17]. Only intestinal schistosomiasis, specifically *S. mansoni*, is prevalent in Uganda and was diagnosed here by duplicate Kato-Katz slides from one stool sample [16]. Detailed socio-demographic, behavioural, household WASH, and environmental data were collected on the same individuals. We fitted a bivariate copula model to separate parasite-specific determinants from residual co-occurrence. From the fitted model we used G-computation to quantify these effects as absolute risk differences and population attributable fractions (PAFs) to estimate how much co-infection each modelled population action could, under strict assumptions, avert. The major aim of this study was to address the following. Which determinants influence both *P. falciparum* and *S. mansoni* versus one of these species, and how much residual dependence remains after accounting for shared determinants?

## Results

There were 4290 participants out of 4330 assessed individuals at the point of recruitment within the SchistoTrack cohort with complete diagnostic and covariate data (Supplementary Fig. S1). The sample composition is shown in Table 1. The prevalence of *S. mansoni* infection was 41.47% (1779/4290) and 41.19% (1767/4290) for *P. falciparum* infection. Co-infection prevalence was 18.51% (794/4290) (Table 1). The RDT (Bioline Malaria Ag *P.f* /Pan) detects *P. fal-ciparum*-specific histidine-rich protein 2 (HRP2) and pan-*Plasmodium* lactate dehydrogenase (pLDH) [28]. Of the 1767 positive tests, 1403 (79.4%) showed the HRP2 line only, 359 (20.3%) both the HRP2 and pLDH lines, and 5 (0.3%) the pLDH line only. Microscopy did not show pLDH-only as *P. falciparum* positive. However, given the very low prevalence of pLDH-only, the possibility that pLDH lines also reflect *P. falciparum*, and the low sensitivity of microscopy, we therefore interpret RDT positivity as *P. falciparum* infection. The two infections were positively associated without adjustment for other variables (*χ*^2^ = 14.87, *p <* 0.001). The crude cross-product odds ratio (OR) was 1.27 (95% confidence interval (CI) 1.13 to 1.44), corresponding to a Yule’s Υ of 0.06 (95% CI 0.03 to 0.09). Given the two marginal prevalences, the expected co-infection prevalence under crude independence was 17.08%, whereas the observed prevalence of 18.51% was slightly higher. Geographically, *S. mansoni* prevalence was highest in Pakwach (45.59%, 802/1759), followed by Buliisa (41.32%, 602/1457) and Mayuge (34.92%, 375/1074).

**Table 1:** Characteristics of participants by infection status. Values are *n* (%) or mean (SD). Household variables apply to all household members. Distances are measured from the home as ln(1+metres). Treatment was self-reported for the year (praziquantel) or month (antimalarials) before recruitment. Water contact is reported weekly activity irrespective of occupation. *p* values are from Kruskal–Wallis or *χ*^2^ tests, with Fisher’s exact test for small counts. *S. mansoni* infection was defined by Kato-Katz and *P. falciparum* infection by RDT. NDVI, normalised difference vegetation index.

|  | Uninfected | *S. mansoni* | *P. falciparum* | Co-infected | Total | *p* |
|---|---|---|---|---|---|---|
|  |  | only | only |  |  |  |
|  | (*n* = 1538) | (*n* = 985) | (*n* = 973) | (*n* = 794) | (*n* = 4290) | value |
| *Individual level* |  |  |  |  |  |  |
| Age (years) | 31.87 (19.40) | 27.36 (16.35) | 18.20 (16.96) | 17.18 (13.00) | 25.01 (18.24) | <0.001 |
| Gender |  |  |  |  |  | <0.001 |
| Male | 575 (37.4) | 435 (44.2) | 488 (50.2) | 458 (57.7) | 1956 (45.6) |  |
| Female | 963 (62.6) | 550 (55.8) | 485 (49.8) | 336 (42.3) | 2334 (54.4) |  |
| Education (years) | 3.67 (3.14) | 3.83 (2.82) | 2.50 (2.44) | 3.13 (2.29) | 3.34 (2.82) | <0.001 |
| Occupation |  |  |  |  |  | <0.001 |
| None/other | 993 (64.6) | 641 (65.1) | 826 (84.9) | 681 (85.8) | 3141 (73.2) |  |
| Farmer | 378 (24.6) | 163 (16.5) | 95 (9.8) | 58 (7.3) | 694 (16.2) |  |
| Fisher | 101 (6.6) | 127 (12.9) | 39 (4.0) | 43 (5.4) | 310 (7.2) |  |
| Fishmonger | 66 (4.3) | 54 (5.5) | 13 (1.3) | 12 (1.5) | 145 (3.4) |  |
| Village majority religion | 1056 (68.7) | 726 (73.7) | 744 (76.5) | 642 (80.9) | 3168 (73.8) | <0.001 |
| Village majority tribe | 1090 (70.9) | 758 (77.0) | 757 (77.8) | 667 (84.0) | 3272 (76.3) | <0.001 |
| Praziquantel, past year | 764 (49.7) | 439 (44.6) | 404 (41.5) | 356 (44.8) | 1963 (45.8) | <0.001 |
| Antimalarial, past month | 201 (13.1) | 112 (11.4) | 218 (22.4) | 115 (14.5) | 646 (15.1) | <0.001 |
| *Water contact (open water)* |  |  |  |  |  |  |
| Fishing | 199 (12.9) | 181 (18.4) | 73 (7.5) | 115 (14.5) | 568 (13.2) | <0.001 |
| Fetching water | 298 (19.4) | 208 (21.1) | 180 (18.5) | 161 (20.3) | 847 (19.7) | 0.494 |
| Washing clothes | 470 (30.6) | 294 (29.8) | 236 (24.3) | 196 (24.7) | 1196 (27.9) | <0.001 |
| Bathing | 65 (4.2) | 60 (6.1) | 67 (6.9) | 47 (5.9) | 239 (5.6) | 0.027 |
| Fishmongering | 98 (6.4) | 64 (6.5) | 18 (1.8) | 17 (2.1) | 197 (4.6) | <0.001 |
| Swimming | 46 (3.0) | 31 (3.1) | 40 (4.1) | 76 (9.6) | 193 (4.5) | <0.001 |
| *Household level* |  |  |  |  |  |  |
| Home quality score | 6.31 (3.55) | 5.88 (3.48) | 5.32 (3.29) | 5.09 (3.16) | 5.76 (3.44) | <0.001 |
| Electricity | 621 (40.4) | 382 (38.8) | 332 (34.1) | 254 (32.0) | 1589 (37.0) | <0.001 |
| Home owned | 1333 (86.7) | 814 (82.6) | 879 (90.3) | 693 (87.3) | 3719 (86.7) | <0.001 |
| Number of rooms |  |  |  |  |  | <0.001 |
| Fewer than 2 | 411 (26.7) | 290 (29.4) | 345 (35.5) | 233 (29.3) | 1279 (29.8) |  |
| 2 or more | 1127 (73.3) | 695 (70.6) | 628 (64.5) | 561 (70.7) | 3011 (70.2) |  |
| Household size | 3.70 (1.48) | 3.65 (1.48) | 3.69 (1.49) | 3.62 (1.37) | 3.67 (1.47) | 0.623 |
| Household social status | 124 (8.1) | 62 (6.3) | 80 (8.2) | 70 (8.8) | 336 (7.8) | 0.203 |
| Handwashing facility | 145 (9.4) | 81 (8.2) | 79 (8.1) | 56 (7.1) | 361 (8.4) | 0.249 |
| Improved sanitation | 904 (58.8) | 599 (60.8) | 541 (55.6) | 426 (53.7) | 2470 (57.6) | 0.009 |
| Treats drinking water | 351 (22.8) | 218 (22.1) | 226 (23.2) | 165 (20.8) | 960 (22.4) | 0.620 |
| Protected water source | 367 (23.9) | 255 (25.9) | 179 (18.4) | 190 (23.9) | 991 (23.1) | <0.001 |
| *Environment and access* |  |  |  |  |  |  |
| Distance to nearest |  |  |  |  |  |  |
| Tap or latrine | 5.36 (1.74) | 5.14 (1.56) | 6.17 (2.08) | 5.81 (1.87) | 5.58 (1.85) | <0.001 |
| Drug shop | 6.20 (1.48) | 6.03 (1.47) | 6.40 (1.32) | 6.32 (1.30) | 6.23 (1.42) | <0.001 |
| Health centre | 7.42 (1.03) | 7.60 (0.83) | 7.67 (0.86) | 7.78 (0.69) | 7.59 (0.90) | <0.001 |
| Water-contact site | 5.77 (1.02) | 5.50 (1.08) | 6.07 (0.95) | 5.76 (1.00) | 5.77 (1.03) | <0.001 |
| NDVI around home | 0.42 (0.15) | 0.39 (0.16) | 0.45 (0.13) | 0.42 (0.15) | 0.42 (0.15) | <0.001 |
| Nearest water-contact site |  |  |  |  |  | <0.001 |
| Lake beach | 609 (39.6) | 440 (44.7) | 222 (22.8) | 227 (28.6) | 1498 (34.9) |  |
| Lake marsh | 462 (30.0) | 299 (30.4) | 304 (31.2) | 246 (31.0) | 1311 (30.6) |  |
| Pond or swamp | 249 (16.2) | 146 (14.8) | 257 (26.4) | 192 (24.2) | 844 (19.7) |  |
| Rice paddy | 24 (1.6) | 9 (0.9) | 16 (1.6) | 5 (0.6) | 54 (1.3) |  |
| River or river marsh | 194 (12.6) | 91 (9.2) | 174 (17.9) | 124 (15.6) | 583 (13.6) |  |
| *Study design* |  |  |  |  |  |  |
| District |  |  |  |  |  | <0.001 |
| Mayuge | 463 (30.1) | 238 (24.2) | 236 (24.3) | 137 (17.3) | 1074 (25.0) |  |
| Buliisa | 586 (38.1) | 370 (37.6) | 269 (27.6) | 232 (29.2) | 1457 (34.0) |  |
| Pakwach | 489 (31.8) | 377 (38.3) | 468 (48.1) | 425 (53.5) | 1759 (41.0) |  |
| Recruitment year |  |  |  |  |  | <0.001 |
| 2022 | 1030 (67.0) | 701 (71.2) | 594 (61.0) | 545 (68.6) | 2870 (66.9) |  |
| 2023 | 324 (21.1) | 118 (12.0) | 333 (34.2) | 182 (22.9) | 957 (22.3) |  |
| 2024 | 184 (12.0) | 166 (16.9) | 46 (4.7) | 67 (8.4) | 463 (10.8) |  |

*P. falciparum* prevalence was highest in Pakwach (50.77%, 893/1759) as well, and similar in both Mayuge (34.73%, 373/1074) and Buliisa (34.39%, 501/1457). Co-infection followed the same district ordering as *S. mansoni*, with prevalence of 24.16% in Pakwach (425/1759), 15.92% in Buliisa (232/1457), and 12.76% in Mayuge (137/1074). Both marginal infection prevalences varied across recruitment years (*p <* 0.001). *S. mansoni* prevalence was 43.41% (1246/2870) in 2022, 31.35% (300/957) in 2023, and 50.32% (233/463) in 2024, whereas *P. falciparum* prevalence was 39.69% (1139/2870), 53.81% (515/957), and 24.41% (113/463), respectively. Co-infection prevalence varied less across years, at 18.99% (545/2870) in 2022, 19.02% (182/957) in 2023, and 14.47% (67/463) in 2024.

### Determinants of malaria and *S. mansoni* infection

Covariate-adjusted age-prevalence curves from the bivariate copula model differed in shape between the two infections (Fig. 1, unadjusted curves are shown in Supplementary Fig. S2).

**Fig. 1:**
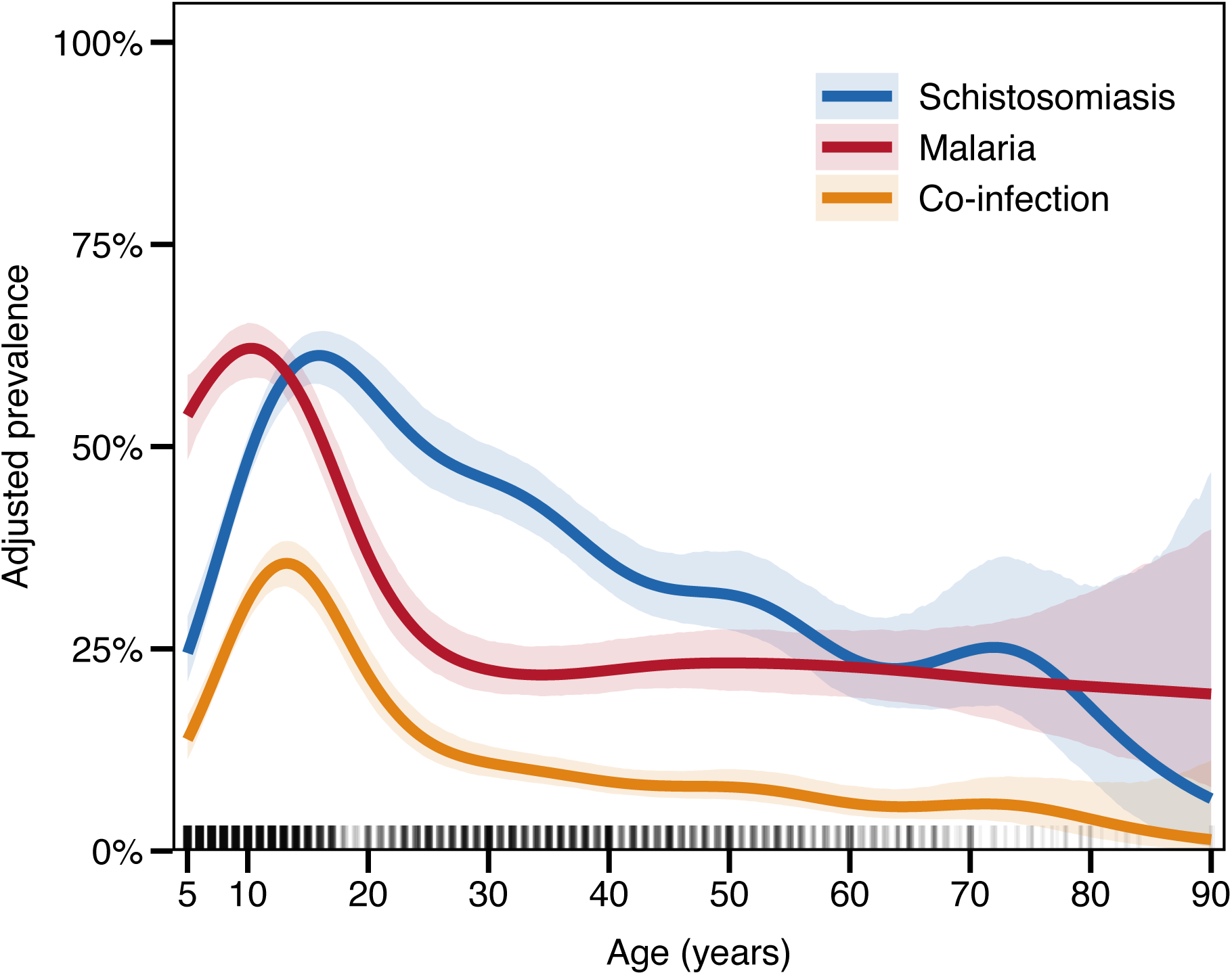
Covariate-adjusted age-prevalence profiles from the bivariate copula model. Penalised splines on age for each model margin and for the joint co-infection probability, with 95% CIs from posterior simulation. Tick marks along the x-axis indicate the age distribution of participants.

*P. falciparum* prevalence peaked at age 10 years (95% CI 9 to 12) and declined gradually through adolescence and early adulthood, levelling off at around 22% from the late twenties.

*S. mansoni* prevalence peaked later, at age 16 years (95% CI 15 to 18), and declined gradually through adulthood. Co-infection prevalence was highest at age 13 (95% CI 12 to 14) in between the two mono-infection peaks. Both age-penalised splines in the *P. falciparum* and *S. man-soni* model margins were non-linear (*p <* 0.001). ORs from the parasite-specific model margins (equations (2) and (3)) are shown in Fig. 2.

**Fig. 2:**
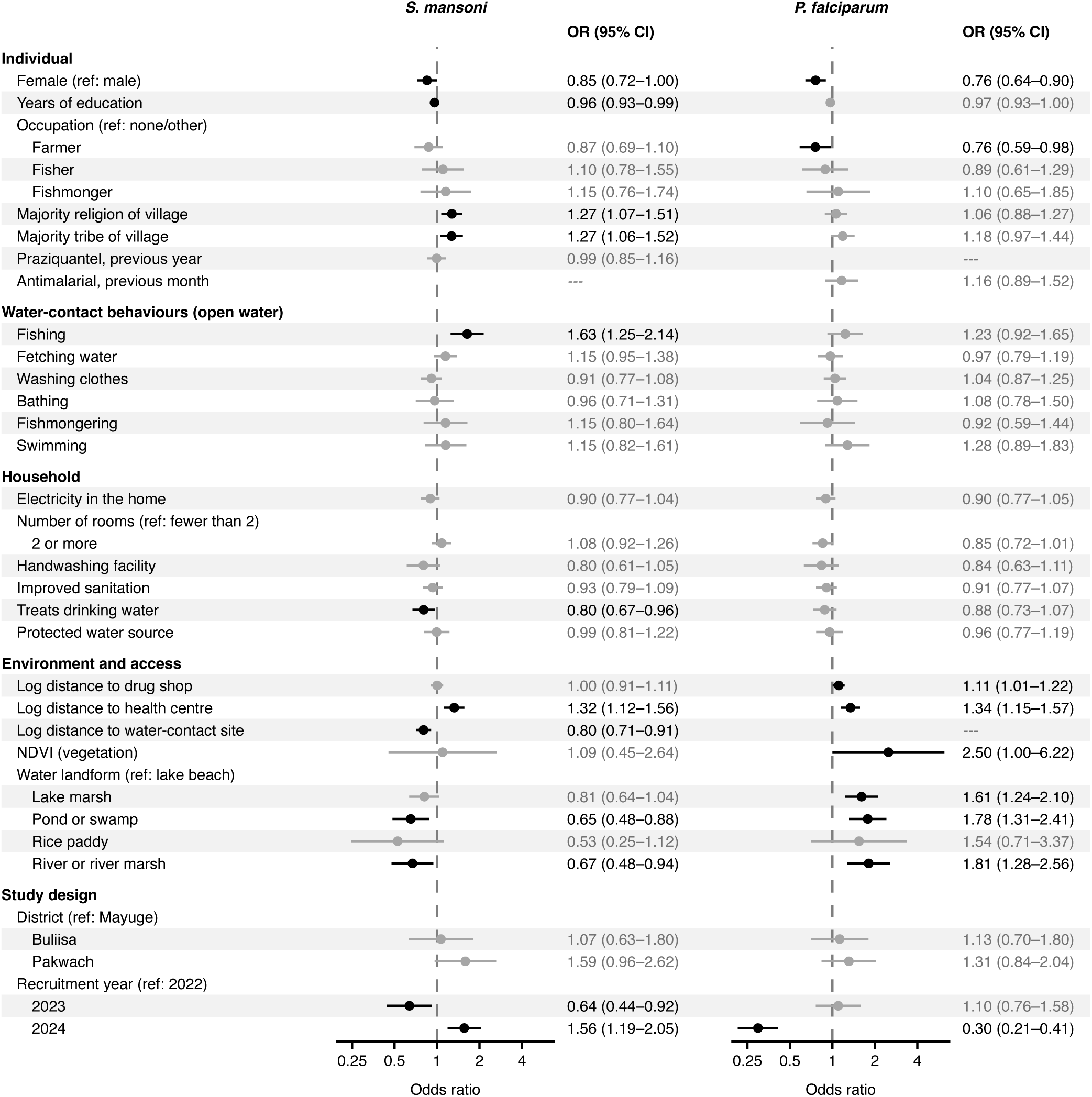
Adjusted odds ratios for *S. mansoni* and *P. falciparum* infection. ORs with 95% CIs from the bivariate copula model. Black points have *p <* 0.05. Dashes mark covariates not included in that model margin. Water contact is reported weekly activity in open water, irrespective of occupation.

In the *S. mansoni* model margin, four factors were associated with higher infection odds. Reported open-water fishing carried the largest of these effects when compared to no reported fishing (OR 1.63, 95% CI 1.25 to 2.14). An individual belonging to the majority religion of their village had higher odds than an individual who did not (OR 1.27, 95% CI 1.07 to 1.51), as did an individual belonging to the majority tribe of their village (OR 1.27, 95% CI 1.06 to 1.52). Odds also increased with distance to the nearest government health centre (OR 1.32 per one-unit increase in log(1+*d*), with *d* in metres; 95% CI 1.12 to 1.56), equivalent to roughly 1.57-fold higher odds at 5 km than at 1 km. Female gender was associated with lower odds of *S. mansoni* infection when compared to male gender (OR 0.85, 95% CI 0.72 to 1.00), and each additional year of education was associated with lower odds of *S. mansoni* infection (OR 0.96, 95% CI 0.93 to 0.99). Distance to the nearest water-contact site showed a similar association, with lower odds among households living farther from that site (OR 0.80 for each one-unit increase in log(1 + *d*), *d* in metres; 95% CI 0.71 to 0.91). The nearest water landform being a pond or swamp (OR 0.65, 95% CI 0.48 to 0.88) or a river or river marsh (OR 0.67, 95% CI 0.48 to 0.94) was associated with lower odds when compared to a lake beach.

In the *P. falciparum* model margin, higher odds were associated with geographic and environmental features. Relative to a lake beach, lake marsh (OR 1.61, 95% CI 1.24 to 2.10), pond or swamp (OR 1.78, 95% CI 1.31 to 2.41), and river or river-marsh (OR 1.81, 95% CI 1.28 to 2.56) all were positively associated with *P. falciparum* infection. Higher vegetation surrounding the home, measured by the normalised difference vegetation index (NDVI), was associated with higher odds of *P. falciparum* infection (OR 2.50 per one-unit increase in NDVI, 95% CI 1.00 to 6.22). The odds of *P. falciparum* infection increased with the distance to the nearest government health centre (OR 1.34 per one-unit increase in log(1 + *d*), 95% CI 1.15 to 1.57), corresponding to roughly 1.61-fold higher odds at 5 km than at 1 km. Female gender was associated with lower odds of *P. falciparum* infection when compared to male gender (OR 0.76, 95% CI 0.64 to 0.90) and individuals who referred to their occupation as farmers had lower odds of *P. falciparum* infection than people who referred to their occupation as none/other (OR 0.76, 95% CI 0.59 to 0.98). No water-contact activity was associated with *P. falciparum* infection when compared to not reporting that activity. Self-reported antimalarial treatment in the month before recruitment was likewise not associated with the odds of *P. falciparum* infection when compared to no reported treatment (OR 1.16, 95% CI 0.89 to 1.52).

### Risk factor classification, effect sizes, and population impact

We report average marginal effects (AMEs) and where a subgroup is named we additionally report conditional AMEs (Figs. 3 and 4). We classify each covariate as convergent, divergent, singular or unclassified according to the sign and sign stability of its two marginal AMEs. We also report PAFs in Table 2 and note that a positive PAF indicates lower predicted prevalence under the given scenario, as a protective factor has a positive PAF when provided for everyone and a negative PAF when removed from everyone.

**Fig. 3:**
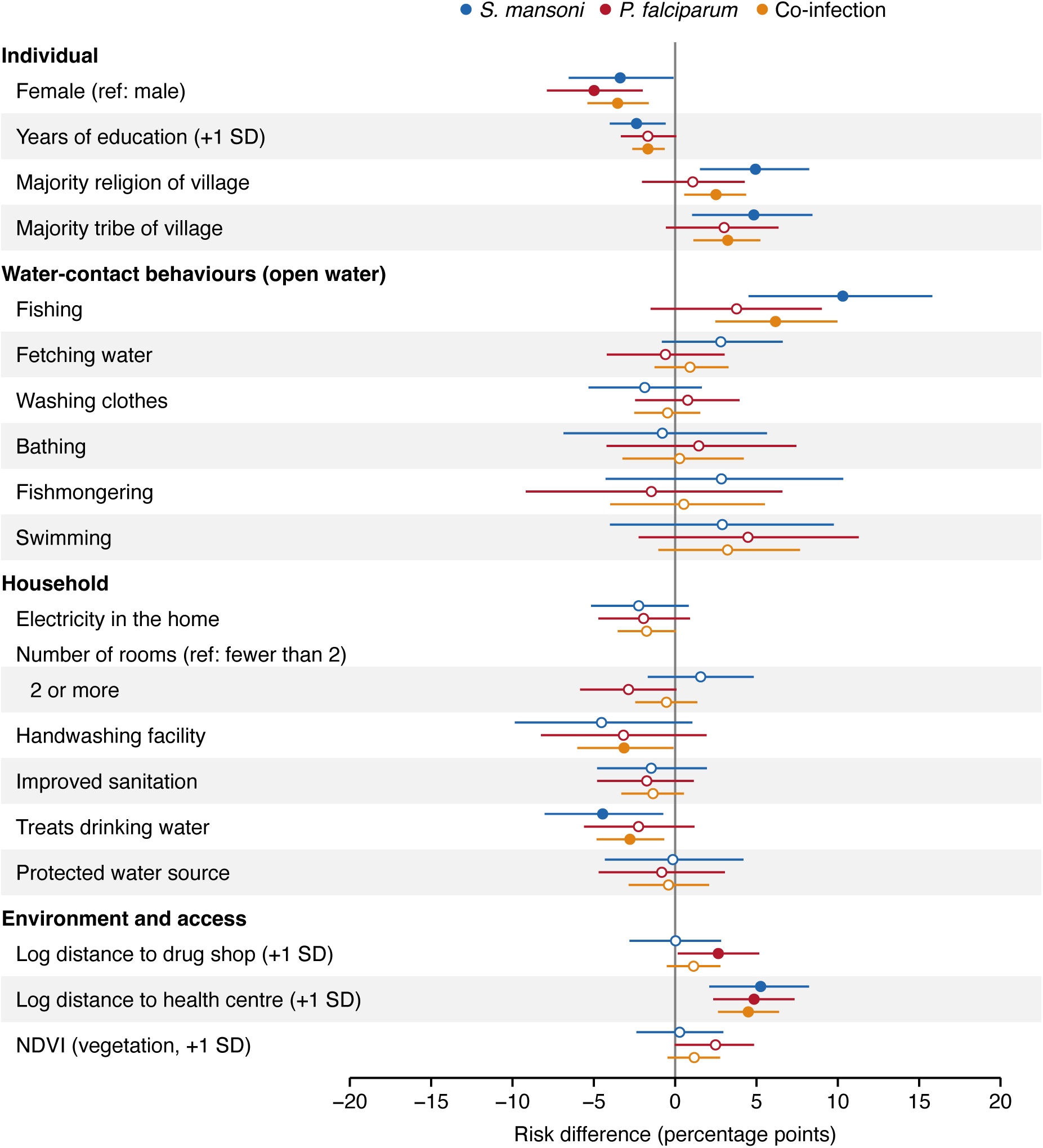
Average marginal effects of covariates included in both model margins. Predicted change in the prevalence of *S. mansoni*, *P. falciparum* and co-infection in percent-age points, with 95% CIs from 5000 posterior draws. Binary covariates compare presence with absence and continuous covariates a one-SD increase. Filled points have CIs excluding zero.

**Fig. 4:**
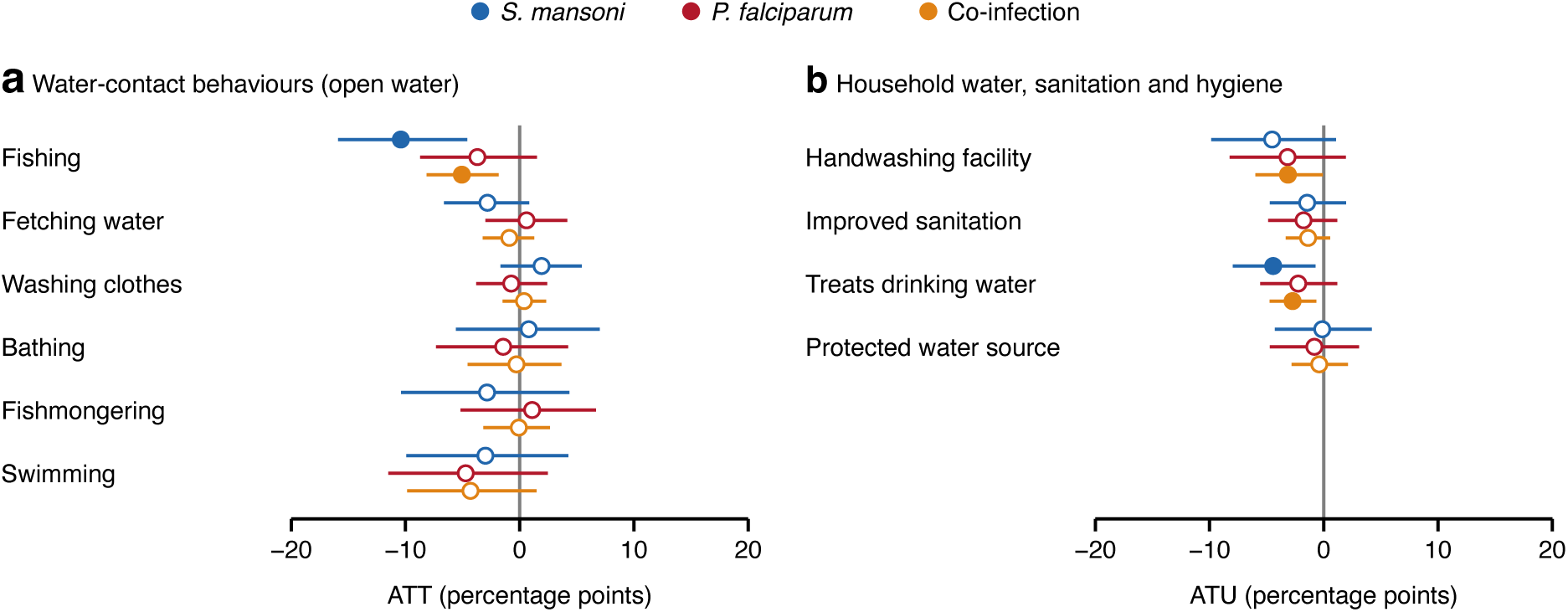
Predicted effects of removing water contact and providing WASH. (**a**) Change in risk if each water-contact behaviour were removed, among participants who reported it. (**b**) Change in risk if each WASH condition were provided, among participants who lacked it. Conditional average marginal effects in percentage points with 95% CIs from 5000 posterior draws. ATT is the average effect among the treated (exposed) and ATU is the average effect among the untreated (unexposed). Filled points have CIs excluding zero.

**Table 2:** Population attributable fractions under modelled scenarios. PAF (%) with 95% CIs from 5000 posterior draws, calculated as (*P*_obs_ − *P*_scenario_)*/P*_obs_. Positive values indicate lower predicted prevalence under the scenario. For individual characteristics and environment, values are standardised contrasts rather than effects of feasible interventions. SD, standard deviation; WASH, water, sanitation and hygiene.

| Variable and scenario | *S. mansoni* | *P. falciparum* | Co-infection |
|---|---|---|---|
| *Individual level* |  |  |  |
| Years of education |  |  |  |
| −1 SD | −5.82 (−9.92, −1.42) | −4.12 (−8.20, 0.17) | −9.58 (−15.44, −3.49) |
| Occupation |  |  |  |
| Set all to none/other | −0.44 (−3.00, 2.21) | −2.06 (−4.41, 0.37) | −2.02 (−4.84, 0.79) |
| *Water contact in open water* |  |  |  |
| Fishing |  |  |  |
| Remove from everyone | 3.32 (1.45, 5.07) | 1.19 (−0.49, 2.81) | 3.63 (1.31, 5.82) |
| Expose everyone | −21.52 (−33.09, −9.34) | −7.98 (−19.15, 3.18) | −29.77 (−47.71, −11.88) |
| Fetching water |  |  |  |
| Remove from everyone | 1.35 (−0.39, 3.16) | −0.28 (−2.01, 1.43) | 0.98 (−1.36, 3.43) |
| Expose everyone | −5.42 (−12.74, 1.57) | 1.17 (−5.90, 8.23) | −3.97 (−14.15, 5.50) |
| Washing clothes |  |  |  |
| Remove from everyone | −1.28 (−3.63, 1.10) | 0.50 (−1.62, 2.58) | −0.57 (−3.44, 2.21) |
| Expose everyone | 3.23 (−2.76, 9.11) | −1.37 (−7.03, 4.34) | 1.95 (−6.06, 10.05) |
| Bathing |  |  |  |
| Remove from everyone | −0.11 (−0.94, 0.74) | 0.20 (−0.57, 0.99) | 0.09 (−1.11, 1.37) |
| Expose everyone | 1.79 (−12.76, 15.66) | −3.33 (−16.90, 9.65) | −1.39 (−21.45, 16.47) |
| Fishmongering |  |  |  |
| Remove from everyone | 0.32 (−0.48, 1.15) | −0.12 (−0.74, 0.57) | 0.02 (−0.65, 0.78) |
| Expose everyone | −6.54 (−23.78, 9.92) | 3.43 (−15.41, 21.36) | −2.85 (−28.61, 20.96) |
| Swimming |  |  |  |
| Remove from everyone | 0.33 (−0.46, 1.08) | 0.52 (−0.27, 1.26) | 1.05 (−0.36, 2.39) |
| Expose everyone | −6.66 (−22.57, 9.17) | −10.35 (−26.05, 5.19) | −16.39 (−38.93, 5.30) |
| *Household level* |  |  |  |
| Electricity |  |  |  |
| Give to everyone | 3.39 (−1.26, 7.82) | 3.00 (−1.41, 7.28) | 6.12 (−0.21, 12.22) |
| Remove from everyone | −2.02 (−4.66, 0.75) | −1.72 (−4.17, 0.81) | −3.38 (−6.82, 0.10) |
| Number of rooms |  |  |  |
| Set all to fewer than 2 | 2.65 (−2.85, 8.14) | −4.83 (−9.85, 0.14) | −2.03 (−9.16, 5.02) |
| Treats drinking water |  |  |  |
| Give to everyone | 8.30 (1.35, 14.91) | 4.23 (−2.22, 10.50) | 11.51 (2.76, 19.85) |
| Remove from everyone | −2.45 (−4.44, −0.40) | −1.24 (−3.08, 0.65) | −3.55 (−6.20, −0.83) |
| Handwashing facility |  |  |  |
| Give to everyone | 9.99 (−2.35, 21.63) | 7.06 (−4.28, 18.40) | 15.58 (0.49, 29.50) |
| Remove from everyone | −0.92 (−2.03, 0.21) | −0.65 (−1.71, 0.40) | −1.43 (−2.80, −0.04) |
| Improved sanitation |  |  |  |
| Give to everyone | 1.49 (−1.98, 4.82) | 1.83 (−1.22, 4.99) | 3.17 (−1.26, 7.61) |
| Remove from everyone | −2.05 (−6.68, 2.69) | −2.41 (−6.60, 1.59) | −4.20 (−10.28, 1.63) |
| Protected water source |  |  |  |
| Give to everyone | 0.27 (−7.76, 7.95) | 1.55 (−5.71, 8.78) | 1.68 (−8.76, 11.80) |
| Remove from everyone | −0.08 (−2.46, 2.36) | −0.45 (−2.59, 1.66) | −0.54 (−3.73, 2.53) |
| *Environment and access* |  |  |  |
| Distance to health centre |  |  |  |
| −1 SD (closer) | 12.27 (4.94, 18.82) | 11.47 (5.60, 17.07) | 21.25 (13.09, 28.62) |
| Distance to drug shop |  |  |  |
| −1 SD (closer) | 0.06 (−6.88, 6.67) | 6.35 (0.39, 12.17) | 6.07 (−2.49, 14.05) |
| NDVI (vegetation) |  |  |  |
| −1 SD | 0.67 (−5.80, 7.00) | 5.94 (−0.06, 11.41) | 6.23 (−2.30, 13.86) |
| Water landform |  |  |  |
| Set all to lake beach | −10.50 (−17.85, −3.07) | 15.51 (8.34, 21.84) | 4.80 (−5.90, 14.50) |
| *Combined scenarios* |  |  |  |
| WASH |  |  |  |
| Give to everyone | 19.61 (5.21, 32.53) | 14.48 (0.82, 27.08) | 29.82 (13.29, 44.03) |
| Remove from everyone | −5.52 (−10.96, 0.13) | −4.78 (−9.90, 0.11) | −9.95 (−17.48, −2.82) |
| WASH without sanitation |  |  |  |
| Give to everyone | 18.22 (3.91, 31.16) | 12.71 (−1.01, 25.11) | 27.24 (10.28, 41.82) |
| Remove from everyone | −3.46 (−6.47, −0.35) | −2.35 (−5.19, 0.50) | −5.61 (−9.70, −1.53) |
| WASH without sanitation or handwashing |  |  |  |
| Give to everyone | 8.56 (−1.52, 18.01) | 5.77 (−3.77, 14.95) | 13.04 (0.17, 25.09) |
| Remove from everyone | −2.53 (−5.49, 0.53) | −1.69 (−4.46, 1.10) | −4.11 (−8.17, 0.02) |
| Water contact |  |  |  |
| Remove from everyone | 3.94 (0.45, 7.34) | 2.01 (−1.19, 5.07) | 5.16 (0.82, 9.19) |
| Expose everyone | −35.37 (−63.57, −3.75) | −18.49 (−48.24, 11.30) | −56.52 (−107.39, −8.08) |
| WASH and water contact |  |  |  |
| Give WASH, remove water contact | 23.39 (8.91, 36.35) | 16.37 (2.35, 29.20) | 33.99 (17.48, 48.02) |
| Remove WASH, expose to water contact | −40.91 (−69.02, −8.97) | −23.39 (−53.26, 6.84) | −68.82 (−122.05, −17.58) |
| WASH without sanitation and water contact |  |  |  |
| Give WASH, remove water contact | 22.03 (7.46, 35.09) | 14.62 (0.53, 27.39) | 31.52 (14.52, 45.74) |
| Remove WASH, expose to water contact | −38.86 (−67.05, −7.15) | −20.90 (−51.02, 9.06) | −63.38 (−115.36, −13.42) |
| WASH without sanitation or handwashing and water contact |  |  |  |
| Give WASH, remove water contact | 12.46 (1.34, 22.35) | 7.74 (−2.56, 17.47) | 17.82 (4.39, 29.86) |
| Remove WASH, expose to water contact | −37.92 (−66.10, −6.28) | −20.23 (−50.18, 9.40) | −61.53 (−113.76, −12.09) |

Distance to the nearest government health centre was the strongest convergent determinant and had the largest estimated association with co-infection. A one-standard-deviation (SD 0.90) increase in log-distance to the nearest government health centre was associated with risk differences of 5.26 percentage points for *S. mansoni* (95% CI 2.10 to 8.23), 4.85 percentage points for

*P. falciparum* (95% CI 2.34 to 7.34), and 4.50 percentage points for co-infection (95% CI 2.63 to 6.39). Moving everyone one standard deviation closer to a government health centre (roughly from 2 km to 0.8 km at the mean log-distance) corresponded to PAFs of 12.27% for *S. man-soni* (95% CI 4.94 to 18.82), 11.47% for *P. falciparum* (95% CI 5.60 to 17.07) and 21.25% for co-infection (95% CI 13.09 to 28.62). Among demographic covariates, female gender was associated with lower risk across all three outcomes compared to male gender, including co-infection (−3.54 percentage points, 95% CI −5.39 to −1.63). Household WASH variables were associated with lower co-infection risk. An individual belonging to a household that treats drinking water had a lower *S. mansoni* risk than an individual belonging to a household that does not (risk difference −4.46 percentage points, 95% CI −8.03 to −0.73; sign stability 0.989), while the *P. falciparum* estimate for the same comparison was in the same direction but not directionally stable (−2.25 percentage points, 95% CI −5.60 to 1.19; sign stability 0.899). The co-infection risk difference was −2.78 percentage points (95% CI −4.83 to −0.67; sign stability 0.995). Among the 3330 individuals living in households that did not treat water, extending water treatment to those households was associated with risk differences of −4.44 percentage points for *S. man-soni* (95% CI −7.97 to −0.73), −2.24 percentage points for *P. falciparum* (95% CI −5.57 to 1.18), and −2.74 percentage points for co-infection (95% CI −4.74 to −0.66; Fig. 4). Extending drinking-water treatment to every individual gave a co-infection PAF of 11.51% (95% CI 2.76 to 19.85). Similarly, among the 3929 individuals living in households without a handwashing facility, providing this facility was associated with risk differences of −4.53 percentage points for *S. mansoni* (95% CI −9.85 to 1.06), −3.17 percentage points for *P. falciparum* (95% CI −8.25 to 1.93), and −3.14 percentage points for co-infection (95% CI −6.00 to −0.10; Fig. 4). Improved sanitation, electricity access and access to a protected water source were unclassified, with neither margin directionally stable (sign stability 0.53 to 0.92; Fig. 3). Water landform was the clearest divergent determinant, with the ORs reported above. Assigning a lake beach as the nearest landform for everyone gave a *S. mansoni* PAF of −10.50% (95% CI −17.85 to −3.07), indicating higher predicted *S. mansoni* prevalence, and a *P. falciparum* PAF of 15.51% (95% CI 8.34 to 21.84), indicating lower predicted *P. falciparum* prevalence. The *S. mansoni* increase under that scenario lines up with the field observations at those sites. Lake beaches held about one third of the snails collected, but 59% of the snails that were shedding cercariae and 46% of the observed sanitation activity (Supplementary Table S1).

Of the six water-contact behaviours reported by individuals, only fishing showed a directionally stable association, limited to the *S. mansoni* margin. The risk difference for reporting fishing in open water was 10.30 percentage points for *S. mansoni* (95% CI 4.51 to 15.81; sign stability 1.000) and 6.17 for co-infection (95% CI 2.47 to 9.98; sign stability 1.000). For *P. falciparum* the estimate was not directionally stable (3.78, 95% CI −1.51 to 9.01; sign stability 0.918, Fig. 4).

Among the 568 individuals who reported fishing in open water, the conditional AME is the same comparison in that subgroup only, and was 10.41 percentage points for *S. mansoni* (95% CI 4.60 to 15.90) and 5.07 percentage points for co-infection (95% CI 1.85 to 8.16). For the scenario of removing all six water-contact behaviours, the co-infection PAF was 5.16% (95% CI 0.82 to 9.19). Providing water treatment, a handwashing facility and a protected water source to everyone, together with no water contact, yielded PAFs of 22.03% for *S. mansoni* (95% CI 7.46 to 35.09), 14.62% for *P. falciparum* (95% CI 0.53 to 27.39) and 31.52% for co-infection (95% CI 14.52 to 45.74). Sanitation had little influence on either parasite, as adding improved sanitation to this scenario changed the PAFs only slightly (23.39%, 16.37% and 33.99%). Excluding handwashing facilities as well roughly halved the co-infection PAF (17.82%, 95% CI 4.39 to 29.86; Table 2).

### Residual co-infection dependence beyond shared determinants

The estimated conditional cross-product OR from the copula model was 1.18 (95% CI 1.01 to 1.39), corresponding to a Yule’s Υ of 0.04 (95% CI 0.00 to 0.08). For malaria status ascertained via RDT, the CI for *ψ* excluded 1, indicating weak positive residual association between the two infections conditional on the determinants included in the marginal equations. The magnitude was small, conditional on the fitted marginal risks. The probability of joint infection was only modestly greater than would be expected under conditional independence. For comparison, the crude cross-product OR was 1.27 and the crude Yule’s Υ was 0.06.

We tested whether *ψ* varied with any single covariate by fitting nine dependence-modulation specifications. None improved on the constant *ψ* baseline by Akaike Information Criterion (AIC) (ΔAIC *≥* 1.86 in every case; Supplementary Table S2). Refitting the full model separately within each district gave *ψ̂* = 1.04 in Mayuge, 1.26 in Buliisa and 1.18 in Pakwach, which were homogeneous on the log scale (Cochran’s *Q* = 0.69, df = 2, *p* = 0.71; Supplementary Table S3).

### Model performance and sensitivity analyses

The pooled 10-fold cross-validated area under the receiver operating characteristic curve (AUC) was 0.712 for *S. mansoni*, 0.786 for *P. falciparum*, and 0.759 for co-infection (Supplementary Fig. S3). Backward elimination dropped five covariates from the candidate set (home ownership, home quality score, household social status, household size, and log-distance to the nearest tap or latrine), in decreasing order of AIC improvement (Supplementary Table S4).

Among the 4290 participants in 2160 households, the household intraclass correlation was 0.039 for *S. mansoni* and was estimated at zero for *P. falciparum*. Refitting the full bivariate model within each cluster-free subsample gave *ψ̂* = 1.23 (95% CI 0.97 to 1.56) among adults and *ψ̂* = 1.20 (95% CI 0.96 to 1.51) among children, both close to the pooled estimate and homogeneous on the log scale (Cochran’s *Q* = 0.02, df = 1, *p* = 0.88; Supplementary Table S5). Adjusted ORs were broadly similar when the model was fitted separately to children aged 5–14 years and to individuals aged 15 years or older (Supplementary Fig. S4). We also compared agreement between RDT and microscopy results and found that they agreed for 80.05% of participants (Cohen’s *κ* = 0.58; Supplementary Table S6), and when refitting the model under alternative malaria outcome definitions requiring detectable parasitaemia, *ψ̂* was 1.06 (95% CI 0.89 to 1.26) and 1.03 (95% CI 0.88 to 1.21), compared to 1.18 under the primary definition (Supplementary Table S7). Replacing field-mapped distance to the nearest water-contact site and water landform with distance to a satellite-derived shoreline gave almost identical discrimination (*S. mansoni* : AUC 0.713, *P. falciparum*: 0.785 and *Co-infection*: 0.759) but poorer fit (ΔAIC +8.6), and *ψ̂* was unchanged at 1.18 (95% CI 1.01 to 1.38; Supplementary Table S8, Supplementary Figs. S5, S6, S7 and S8). Additionally, we computed E-values for the significant marginal associations to quantify robustness to unmeasured confounding (Supplementary Table S9). Excluding recruitment year, E-values for the point estimates of significant associations ranged from 1.17 to 2.54, and E-values for the 95% confidence limit closest to the null ranged from 1.03 to 1.55. The largest were for NDVI and water landform on *P. falciparum*, where an unmeasured confounder would need roughly two-fold associations with both exposure and outcome to explain the estimate. Fishing and distance to health centre were moderately robust (E-values 1.57 to 1.88). We also examined infection intensity and haemoglobin and anaemia data to assess whether our findings extend beyond infection status as a clinical validation (Supplementary Tables S10, S11 and S12, Supplementary Figs. S9, S10, S11 and S12).

## Discussion

Integration of malaria and schistosomiasis control has been advocated for two decades and is reinforced by the WHO roadmap for neglected tropical diseases [6, 7, 8, 2]. Whether control can be jointly targeted depends in part on whether the same individuals are at risk of both infections, and thus on the extent to which *P. falciparum* and *S. mansoni* share individual-level determinants rather than merely co-occur geographically. We examined the shared and parasite-specific determinants of *P. falciparum* and *S. mansoni* infection in 4290 individuals in rural Uganda using a bivariate copula model that estimates each parasite model margin with its own covariate set, evaluates the direction of each effect on both model margins, and quantifies the residual dependence between the two infections in a single fit. We found that the two infections share several upstream determinants whilst retaining parasite-specific risk factors for infection acquisition. Residual dependence between the two infections after inclusion of easily observable covariates was weak.

The burden of both infections was strongly age-dependent with age being an important shared determinant. *P. falciparum* prevalence peaked at 10 years of age and *S. mansoni* prevalence at 16 years of age. The early *P. falciparum* peak is consistent with evidence that school-aged children retain high susceptibility and act as a community transmission reservoir [17, 29], while parasitaemia was highest at 5 years of age, consistent with immunity acquired through repeated exposure [30]. Co-infection prevalence peaked at 13 years of age between the two parasite-specific peaks. Given the weak residual dependence between the two infections, this co-infection age peak is expected if co-infection largely reflects the overlap of two separate risks. Despite differences in the exact age year of peak prevalence, both parasites overlapped substantially within school-aged children. Notably, a substantial burden of both infections remained in adults, who are less often the focus of control programmes [19, 23].

The age patterns we observed have implications for mass treatment campaigns. Both infections and co-infection were most common among school-aged children. Praziquantel has been delivered as preventive chemotherapy through school and community-based MDA in the districts of this study since 2003 [23, 16]. Malaria chemoprevention in school-aged children has been shown to reduce infection prevalence [29], resulting in revised WHO guidance that has widened the age groups eligible for malaria chemoprevention [19] as well as updates to the Uganda national malaria plan to extend community case management for malaria to all age groups [22]. Adding malaria chemoprevention to school-based MDA or providing praziquantel through primary healthcare facilities where antimalarials are distributed requires further research. Realising any integration strategy will depend on platforms that are already under strain, as community case management has been limited by drug stock-outs and uneven coverage across regions [31] and praziquantel distribution through MDA has been interrupted due to the loss of international aid. Additional benefits of treatment integration may include specific effects of artemisinin-based treatments against juvenile schistosomes that are not affected by praziquantel [32]. Both parasites having reservoirs in the adult population and need treatment beyond where the peak prevalence is observed in school-aged children [16, 17]. Other non-modifiable participant aspects such as male gender and living in a western district were shared risk factors for both infections; this information could be used in addition to age to build profiles of individuals requiring targeted treatment or other interventions.

Female gender was associated with lower risk of both infections. Given that the two parasites are acquired through different transmission routes, the effect of shared gender may plausibly be the sum of parasite-specific exposures rather than a single shared behaviour. For *S. mansoni*, the male excess risk is consistent with occupational freshwater contact, as fishing is almost exclusively undertaken by men in these communities and was associated with *S. mansoni* infection in our data [16]. By contrast, none of the water-contact activities we measured were associated with *P. falciparum* infection, so the lower *P. falciparum* risk in females cannot be attributed to the same domestic versus occupational division of water contact. The female risk for *P. fal-ciparum* infection might reflect other exposures, such as outdoor activity during the evening hours when *Anopheles* bite, or differences in care-seeking [33, 34]. Males therefore appear to be a priority group for both infections, and because they attend health facilities less often for routine or preventive care [33, 34], reaching them may depend on community-based rather than facility-based delivery.

Using our bivariate modelling framework, we were able to identify that improved access to primary healthcare services could jointly benefit malaria and schistosomiasis control efforts, thereby offering an opportunity for integration. Greater distance to a government health centre was associated with higher risk of both infections, and reducing log-distance by one standard deviation resulted in the largest single-covariate co-infection PAF observed, at 21.25%, alongside PAFs of 12.27% for *S. mansoni* and 11.47% for *P. falciparum*. Antimalarials reach individuals both at health centres and door-to-door through community case management, whereas praziquantel is not available through routine health centres and is administered only through MDA, as donated tablets are allocated to vertical campaigns delivered by teachers and community medicine distributors rather than government health workers [35]. When campaigns are delayed, communities along Lake Albert therefore seek treatment from private drug shops, village health team members, or traditional healers [36]. The association of distance with *S. mansoni* infection is unlikely to reflect access to treatment and more plausibly captures the remoteness of households that campaigns are more likely to miss [34, 37]. For *P. falciparum*, by contrast, antimalarials are available at health centres, so greater distance may plausibly hinder access. WHO recommends that all infected individuals, regardless of age, be treated with praziquantel at health facilities [23]. Making praziquantel available in primary healthcare facilities would provide an accessible route to treatment between MDA rounds and allow co-infected individuals to be treated for both infections at a single visit. Our findings add population-level support for this recommendation, and we would expect the greatest gains for schistosomiasis, which currently has no routine treatment route within local health systems. Health workers in our study districts were often uncertain about praziquantel dosing outside MDA, and called for its inclusion on the national essential medicines list [35]. Additional research is needed to evaluate whether stocking praziquantel in health centres, together with training in diagnosis and dosing, increases treatment coverage for both infections, and whether managing both infections at the same facility reduces co-infection.

We found that no single WASH component consistently lowered the risk of both infections. However, providing drinking-water treatment, handwashing facilities and access to protected water sources, together with no water contact, could reduce co-infection by almost a third (31.52%). WASH could therefore serve as a shared platform for both programmes. For schistosomiasis, the benefit is well established, as water supplies reduce contact with open water, and high community coverage may protect even those without household access [38]. Households that adopt WASH practices may also use bed nets and live in better housing, which is itself associated with lower malaria risk [39], although the housing quality score we considered did not improve our model. A household WASH package of this kind would be a complex intervention, but comparable approaches have succeeded for both infections. An integrated strategy in China combining safe water, sanitation, health education and the removal of cattle from snail habitats reduced *Schistosoma japonicum* prevalence to below 1% [40], and environmental management, including drainage and housing modification, has substantially reduced malaria morbidity and mortality [41]. Efforts to integrate the two programmes have so far focused on co-administering praziquantel and antimalarials or on health education, and few studies of co-infection have considered WASH at all [9, 12]. Eventually there is a need to test complex household WASH interventions in randomised trials for reducing *P. falciparum* and *S. mansoni* co-infections to identify whether such interventions are a valid approach for integrated control of malaria and schistosomiasis.

Any water contact has been proposed to increase the odds of co-infection [12, 21]. However, we found that fishing in open water was the only water-contact activity associated with infection, and only with *S. mansoni*, even though fishing sites in marshy areas also provide habitats suitable for mosquitoes [18, 42]. Policymakers in the future will be confronted with open questions about which interventions can be integrated and where parasite-specific control remains necessary. Our framework separates determinants that act on both infections, from those that act on one parasite, such as fishing and the type of water site, which require targeted measures. Reducing *S. mansoni* infection at water sites, through focal snail control at frequently visited sites near homes [43] and praziquantel treatment of the people who use them, would also prevent co-infection, but would not reduce malaria. Such measures should therefore be planned with malaria in mind, as changes at water sites can create or remove mosquito breeding sites [41]. In agricultural and irrigation areas, integrated vector management combining environmental management with chemical and drug-based measures, including mollusciciding and larval source management, has been a successful approach to controlling both mosquito and snail-borne diseases [44]. Future research should test whether snail control and mosquito larval source management can be delivered jointly at the same water sites, and whether schistosomiasis measures can be designed to also reduce mosquito breeding or, at a minimum, avoid creating new breeding sites.

For occupations, only farming was associated with either infection, with lower *P. falciparum* risk than in individuals without occupation and no difference in *S. mansoni* infection risk, consistent with earlier findings in this cohort [17]. Occupational water contact rises among males from adolescence onwards [16], so those at highest risk fall outside school-based praziquantel programmes [23]. Community-wide treatment in fishing communities, for example at landing sites, should therefore reach everyone who fishes, and could also offer malaria testing and treatment. Combined delivery of this kind has precedent. When bed nets and malaria treatment were added to the work of community ivermectin distributors, coverage of both more than doubled [45]. The farming category used in our analysis, however, combined subsistence farmers with rice farmers and papyrus gatherers (given the few individuals in the latter categories), whose work in standing water and wetlands could expose them to both parasites. Future studies should examine these occupations separately and test whether combined outreach at landing sites can deliver treatment for both infections to adults.

The landform of the nearest water site of a household was the clearest divergent determinant. Compared to lake beaches, ponds, swamps, and rivers and river marshes were associated with lower *S. mansoni* but higher *P. falciparum* risk. The two parasites have distinct niches, and one cannot be inferred from the other, which makes their ecology difficult to target jointly [18, 14]. Knowing where exposure occurs for each parasite therefore remains a prerequisite for targeting, even where the ecology is not shared. Distance to the nearest water-contact site is central to locating schistosome exposure but far less relevant to malaria [43]. In this dataset, satellite-derived shoreline distance performed similarly to field-mapped distance in the *S. mansoni* model, suggesting that it may serve as a suitable proxy indication in comparable lakeside settings. The satellite-derived shoreline measure could extend the field-mapped water-site component of the *S. mansoni* margin to unsurveyed parts of the Lake Albert and Lake Victoria basins, using freely available Sentinel-2 imagery [46]. This approach may offer a scalable source of parasite-specific ecological information that could be used in programme planning.

Through our descriptive analysis, we found that co-infected individuals did not necessarily have higher parasite loads. Co-infected individuals carried modestly higher *S. mansoni* egg counts than those with schistosome infection alone, but lower *P. falciparum* parasitaemia. This difference was limited to those with the heaviest schistosome infection. Community-based paediatric cohorts in Mali and Senegal found low-to-moderate *Schistosoma* egg counts were associated with lower parasitaemia and fewer febrile episodes [10, 47, 48], with reduced regulatory T-cell responses and dampened Th1 cytokine production proposed as the mechanism [49]. It is unclear whether there is a biological interaction to explain the results in our study, i.e. the lack of heavy co-infection clustering, as we did not have immunological data and this area would require additional studies. From a clinical perspective, if co-infected individuals carried a greater burden of anaemia, its management would need to account for co-infection [24, 25]. We found little evidence for this, as anaemia associated with infection seemed to be primarily due to *P. falciparum*. Co-infected individuals were no more anaemic than those with *P. falciparum* alone and *S. man-soni* mono-infection accounted for only 0.8% of anaemia. There was no interaction between the two infections on haemoglobin or on the additive scale for anaemia. Hookworm, a common cause of anaemia, was uncommon outside Mayuge, and adjusting for it in this analysis did not change the estimates.

Our analysis addresses a methodological gap identified in recent systematic reviews of co-infection [13, 12]. All 30 studies in our recent review of malaria and schistosomiasis co-infection collapsed the four infection states into a single binary outcome, which obscures through which infection a determinant acts [12]. Our framework models the two infections as separate margins coupled through a Plackett copula, with a dependence parameter that captures whether co-infection is more common than expected given each infection risk [50, 51]. This framework allows us to identify which determinants are shared and which are parasite-specific, and thereby to distinguish the measures that a joint programme can deliver from those each individual programme must retain, which will become crucial for integration efforts advocated for by the WHO.

The empirical strengths of this study are its scale, wide age range, and diverse geographic setting within rural Uganda. Methodologically, the copula model proposed here is able to return marginal effects, residual dependence, and joint co-infection probabilities in a single fit; the G-computation approach keeps the projected scenarios internally consistent with the fitted margins. Several limitations should be noted. The cross-sectional design cannot establish temporal ordering of exposures or distinguish prevalent from incident infection, and because recruitment took place in January and February, we may have captured malaria near its seasonal high and *S. mansoni* below its seasonal peak [52, 53]. We did not adjust for bed-net use because it was not recorded for participants recruited in 2022, two thirds of the sample, so the associations of house-hold WASH may partly reflect differences in bed-net use, although unmeasured shared exposures of this kind would also be expected to appear as residual dependence, which was weak. Individuals were paired within household, but residual household clustering was negligible (design effect 1.04 for *S. mansoni* and 1.00 for *P. falciparum*) and estimates were unchanged in cluster-free subsamples. Residual dependence was smaller with malaria defined based on detectable parasitaemia, so the primary estimate is the largest of those considered. The projected PAFs depend on assumptions of conditional exchangeability, positivity, consistency, and correct model specification that cannot be verified in observational data, so the combined scenario should be read as a ceiling and not a forecast [54]. Finally, the cross-product OR used to summarise residual dependence is non-collapsible, so future comparisons across populations or specifications must separate differences in dependence from aggregation artefacts [55].

*P. falciparum* and *S. mansoni* can be targeted through easily observable shared and parasite-specific risk factors. Age, gender, district of residence, access to primary healthcare services, and complex WASH interventions show promise for targeting individuals through shared treatment campaigns and for developing future integrated interventions to prevent co-infections. We provide a generalisable framework and comprehensive evaluation of shared risk for malaria and schistosomiasis. If our findings are replicated in other geographic settings, the shared risk factors identified here may inform the targeting and rollout of large-scale interventions for the integration of treatment and control for malaria and schistosomiasis.

## Methods

### Study design and population

We conducted a cross-sectional study using data from all participants at the point of recruitment into SchistoTrack, a community-based prospective cohort in Uganda established in 2022 [26, 27]. The cohort is in three districts: Pakwach along the River Nile and Lake Albert, Buliisa along Lake Albert, and Mayuge along Lake Victoria. Households were randomly sampled from local village registers across 52 villages. Recruitment and data collection took place in January and February of each study year (2022, 2023, 2024), during the dry season following the high-transmission rainy period [52]. Households were eligible if they included at least one child and one adult, who had resided in the community for at least six months of the year. One adult (aged *≥* 18 years) and one child (aged 5 to *<* 18 years) per household were enrolled using a paired sampling approach. Comparison of the clinical cohort against the full household census has shown no demographic bias with respect to age or gender [26]. The final analytic sample included 4290 individuals, after removing individuals with missing diagnostic results for either *P. falciparum* or *S. mansoni* and incomplete covariate data. Each participant contributed a single observation.

### Outcome ascertainment

*P. falciparum* infection was ascertained by RDT (Bioline Malaria Ag *P.f* /Pan), which detects *P. falciparum* histidine-rich protein 2 (HRP2) and pan-*Plasmodium* lactate dehydrogenase (pLDH), on a finger-prick blood sample [28]. We encoded positive RDT results hereafter with *Y_M_*= 1 for a positive result and *Y_M_*= 0 otherwise. Parasitaemia intensity was assessed by thick and thin blood film microscopy following WHO guidelines [19].

*S. mansoni* infection was confirmed via Kato-Katz stool microscopy on duplicate slides prepared from one stool sample [56]. Infection was coded as *Y_S_* = 1 if EPG exceeded zero on any slide and *Y_S_* = 0 otherwise. We retained the infection intensity data of both infections for descriptive comparisons between mono- and co-infected individuals, although they were not used in the primary bivariate analysis for which the main outcome was the pair (*Y_S_, Y_M_* ) *∈ {*0, 1*}*^2^. As secondary outcomes we compared infection intensity between mono- and co-infected individuals as well as altitude-adjusted haemoglobin and anaemia across the four infection groups, for which we also considered an interaction between the two infections.

### Covariates

At the individual level we included age (years), self-reported gender (male as reference), years of formal education (0 to 14), and occupation coded as a four-level categorical variable including none/other (including students and unemployed), farmers, fishers, and fishmongers. The none/other category was chosen as the reference category for analysis [20, 26]. Occupation is defined as the primary income-generating activity. Additionally, binary indicators recorded whether each individual belonged to the majority tribe or majority religion within their village [20].

We additionally considered six water-contact behaviours in both marginal models as binary indicators and included fishing, swimming, fetching water from open water bodies, washing clothes in open water, fishmongering at water sites, and bathing in open water. These indicators were also previously used in an analysis by Reitzug et al. [16] focusing on the association between water contact and schistosomiasis. The behaviours were recorded by structured household interviews with the household head or spouse as a respondent for each household member, and referred to typical weekly activity, as described by Reitzug et al. [16]. All water contact indicators record activity irrespective of occupation, as individuals engage in activities such as fishing without it being their main source of income.

Household-level variables were as follows. To account for socio-economic status we included a composite home-quality score (3 to 12 scale) based on wall material, roof type, and floor quality used in previous analysis [20, 17]. We also considered access to electricity, home ownership, number of rooms, and household size. Household social status was coded as positive if any member held a recognised local leadership position.

Since WASH access may affect infection of either parasite, we assessed at the household level the availability of a handwashing facility with soap and water, sanitation facility type, classified as improved or unimproved per WHO/UNICEF Joint Monitoring Programme criteria [57], any water-treatment practice (bleach or chlorine, solar disinfection, or cloth straining) and access to a protected water source (tap or borehole).

Based on infrastructure field surveys in 2022 and 2023, health facility locations were available, allowing us to calculate the Euclidean distance from each household to the nearest government health centre and to the nearest private drug shop. For modelling purposes we applied to both variables a log transformation and included them in the model as log(1+*d*) with *d* in metres [26]. Additionally, we included binary indicators of self-reported praziquantel treatment in the preceding year before recruitment and self-reported antimalarial treatment during the month before recruitment in the respective model margins.

To capture environmental exposure relevant for transmission of both parasites, we derived the NDVI from Sentinel-2 Level-2A surface reflectance imagery, which was further processed by hexagonal grid aggregation and Gaussian-weighted neighbourhood smoothing as described in Lang et al. [17]. We also included the water-body type of the nearest water-contact site (lake beach, lake marsh, river or river marsh, pond or swamp, rice paddy) to capture ecological features that may influence transmission [18]. In the *S. mansoni* margin we also considered the distance from the household to the nearest water-contact site as a spatial covariate. We also included district (Mayuge, Buliisa, Pakwach) and recruitment year (2022 to 2024) in both margins to account for unobserved differences between study sites and recruitment years [26]. Because the two parasites have distinct transmission routes, the marginal models use overlapping but not identical covariate subsets.

### Measure of residual dependence

We summarised dependence between *S. mansoni* and *P. falciparum* infection using the cross-product OR as the measure of association. Co-infection of two parasites in the same individual may arise from shared exposure or upstream covariates, from residual association that persists after those determinants are accounted for, or from a mix of both. In observational data, crude co-infection prevalence does not distinguish these two sources. We therefore require a measure of association that does not change with the marginal prevalences of either infection, so that comparisons across populations or model specifications can capture whether co-infection is more or less likely than by chance given the marginal prevalences. For two binary outcomes, the joint distribution is characterised by the two marginal probabilities and one additional degree of freedom, which can be captured by the cross-product OR

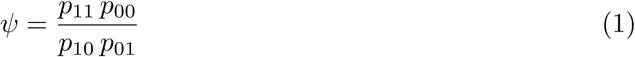

in our model. We used *ψ* because it is margin-free, i.e. it is invariant under independent rescaling of rows or columns, and therefore does not change with the prevalence of either infection. Any other margin-free measure for a 2 *×* 2 table is additionally a monotonic function of *ψ* [51].

Independence corresponds to *ψ* = 1 and values above 1 indicate that the two infections co-occurred more often than the marginal prevalences would imply. Because *ψ* is unbounded, we also report Yule’s coefficient of colligation, 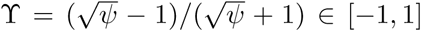, as a bounded summary [51]. We write it as Υ rather than *Y* , following Geenens [51], to avoid confusion with the outcome variables *Y_S_* and *Y_M_* . Υ equals the Pearson correlation of the 2 *×* 2 table after both margins have been standardised to 0.5, and is therefore likewise invariant to the prevalence of either infection. Since 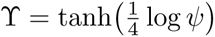 is strictly increasing in *ψ*, intervals for Υ were obtained by transforming the boundaries of the interval for *ψ*.

### Bivariate copula model

In the following, we denote the covariate vectors entering the *S. mansoni* model margin as **x***_S,i_* and as **x***_M,i_* for the malaria model margin for an individual *i*, and let *v*(*i*) index the village. The marginal logits are

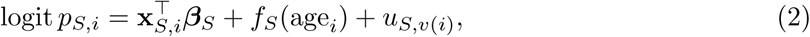

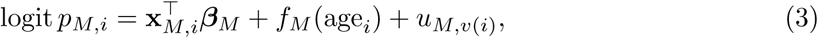

where *u_S,v_* and *u_M,v_* denote village-level random intercepts. We modelled age with a penalised spline in each margin, as the relationship of each infection with age is non-linear and differs in shape between the two parasites. For *S. mansoni*, *f_S_* is a thin-plate regression spline [58]. For *P. falciparum*, *f_M_* is an adaptive spline, a penalised spline with a smoothing penalty that itself varies with age, with an order-3 penalty as in our previous analysis of this cohort [17]. For comparison, unadjusted age-prevalence curves for *S. mansoni*, *P. falciparum* and co-infection were each estimated with a logistic generalised additive model (GAM) containing only a penalised spline of age [58] (Supplementary Fig. S2). We link both margins through a copula to obtain the joint co-infection probability

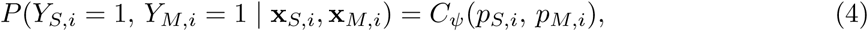

where *C_ψ_* denotes the closed-form Plackett formula parameterised by the cross-product OR *ψ* defined in equation (1) [51]. Algebraically, this expression is identical to the reconstruction of a 2 *×* 2 table from its margins and its OR. A continuous copula family is not identifiable from two binary outcomes, therefore the Plackett copula here denotes a convenient odds-ratio parameterisation of the bivariate Bernoulli distribution. The closed-form expression, its derivation, and further discussion are provided in Supplementary Note 1.

We estimated the model by penalised maximum likelihood jointly with the GJRM package (version 0.2.6) in R (version 4.2.1) [59], with the smoothing parameters selected automatically by minimising an approximate AIC within the fitting algorithm. Our analysis operates within an empirical Bayes framework that links penalised likelihood and Bayesian smoothing [60, 58], such that each penalty term is equivalent to a Gaussian prior distribution on the corresponding coefficients, 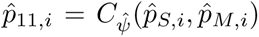 is a (pseudo-)inverse of the penalty matrix [60]. The penalised estimate ***β̂*** is the corresponding maximum a posteriori estimate and instead of assigning the smoothing parameters hyper-prior distributions, we estimated them directly from the data [60, 58]. Conditional on the estimated smoothing parameters, the coefficients then have the approximate Gaussian posterior ***β*** *|* **y** *∼ N* (***β̂***, **V***_b_*) from which intervals approximately attain nominal frequentist coverage across the function [61, 59].

We drew *B* = 5000 samples from the posterior of the fitted coefficients [58] and evaluated each estimand at every draw. For AMEs, PAFs and the age-prevalence curves, the 95% intervals are the 2.5th and 97.5th percentiles of the draws (Supplementary Note 1). ORs and *ψ* have Wald intervals from **V***_b_*. Given the close nominal frequentist coverage we report them as 95% confidence intervals (CIs) throughout [61]. All tests were two-sided.

We assessed out-of-sample discrimination by 10-fold cross-validation, during which the folds were stratified on the cross-classified schistosomiasis-by-malaria outcome, so that each fold carried a comparable share of co-infected individuals. Within each fold the full model, including age splines and village random intercepts, was refitted on the nine training folds and from the refitted model the infection probabilities for *S. mansoni* and *P. falciparum* for the held-out observations were obtained. We pooled the predictions across folds and computed a single AUC. We obtained classification thresholds using Youden’s *J* in the training folds only, so that no held-out observation contributed to its own threshold [62].

### Covariate selection and model evaluation

Covariates were included in the marginal models based on prior work in this cohort [16, 17]. We retained a locked subset irrespective of model fit which included age, district, recruitment year, gender, occupation, education years, praziquantel or antimalarial treatment status, the six water-contact behaviours, the four WASH variables, electricity access, and the spatial covariates of primary interest (mean NDVI, log-distance to nearest private drug shop, log-distance to nearest health centre). The remaining covariates, listed in Supplementary Table S4, were selected via joint backward elimination on the bivariate model. Inference on the retained terms depends on the selection path, although the extent of this concern is limited by the pre-specified set of covariates.

### Risk differences and population attributable fractions

From the fitted model we obtain three predicted probabilities for each individual. A marginal *S. mansoni* risk *p̂_S,i_* estimate, marginal *P. falciparum* risk *p̂_M,i_* estimate, and joint co-infection risk estimate *p̂*_11*,i*_ = *C_ψ̂_*(*p̂_S,i_, p̂_M,i_*). From these estimates we derived standardised risk differences via G-computation [54]. For each binary covariate *Z*, we generated two scenario datasets in which *Z* was set to 1 and to 0 for every individual while all other covariates were held at their observed values, to obtain the predicted infection probabilities from the model. For each setting these predicted probabilities were averaged across the population and then differenced. For continuous covariates, the scenario was a one-standard-deviation increase from each individual’s observed value. We provided full definitions in Supplementary Note 1 and emphasise that these estimates rest on conditional exchangeability, positivity, consistency, and correct specification of the outcome model [54].

We report the AME over the full sample, the PAF, and the conditional AME among the relevant subgroup (currently exposed for the six water-contact behaviours, currently unexposed for the four WASH variables). Under the stated assumptions, the conditional AME among the unexposed quantifies the predicted risk reduction if an intervention were extended to those who currently do not have access to it. Each covariate was classified according to the signs of its two marginal AMEs and the stability of those signs across the posterior draws. We calculated the sign stability as the proportion of the *B* = 5000 draws in which an AME takes the same sign as its point estimate, ranging from 0.5 to 1, and considered a margin as directionally stable when sign stability was at least 0.975. A covariate was convergent if both margins were directionally stable and agreed in sign, divergent if both were directionally stable with opposite signs, singular if exactly one was directionally stable, and unclassified if neither was. Multi-level factors and intervention groups were classified by the same rule applied to setting all individuals to the reference level or to a specific scenario. Full definitions are in Supplementary Note 1.

We also computed joint PAFs for three blocks of covariates. The three blocks included changing all four WASH variables together to either full coverage or the reference level for individuals (WASH block), changing all water-contact variables together (water-contact block), and a combined block between the two. The combined block paired full WASH coverage with no measured water contact, and its reverse paired no WASH with universal water contact. We repeated the WASH and combined blocks excluding sanitation, and excluding both sanitation and handwashing facilities.

### Sensitivity analyses

We conducted six sensitivity analyses. First, we replaced the field-mapped distance to the nearest water-contact site with the satellite-derived shoreline distance and removed the field-derived water-body type variable, since both depend on the same site mapping. Our goal was to assess a model specification that requires no field-based snail or water-contact site data, that we can compare to the main model. We derived water bodies from Sentinel-2 imagery using the Modified Normalised Difference Water Index on the same hexagonal grid as the NDVI [63, 17, 46], classified water and land by Otsu’s threshold [64], and defined shoreline hexagons as those adjacent to the opposite class. Distance to the nearest shoreline hexagon entered the *S. mansoni* margin as log(1 + *d*). The entire pipeline and its validation against field-mapped sites are described in Supplementary Note 2 and Supplementary Fig. S6. Second, we refit the full bivariate model separately within each district, allowing both margins and the dependence parameter to vary, to describe district-specific residual dependence (Supplementary Table S3). We also evaluated the homogeneity by comparing the constant-*ψ* baseline to a model allowing log *ψ* to vary by district via likelihood ratio test, holding both margins common so that the comparison isolates dependence. We also report pairwise Wald contrasts on the log scale in Supplementary Table S3 as a descriptive summary of heterogeneity. Third, to test whether residual dependence concentrates in particular subgroups, we modelled *ψ* as a function of covariates on a third linear predictor, 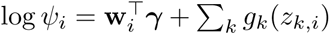, where **w***_i_* is a low-dimensional subset of the marginal covariates and *g_k_* are penalised splines, which we specified with nine differing choices of **w***_i_* (age, gender, district, log-distance to the nearest water-contact site, and combinations of these). We compared these specifications against the constant-*ψ* baseline by AIC, for which the full specifications and results can be found in Supplementary Table S2. Fourth, because each household contributed one adult and one child, individuals were paired within household and all household-level covariates were shared within pairs. We quantified residual clustering by fitting intercept-only mixed-effects logistic models with village and nested household random intercepts to each infection separately, and computed the household intraclass correlation on the latent scale 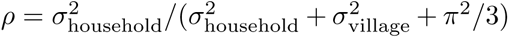. Additionally, we refitted the full bivariate model separately among adults and among children. Fifth, because HRP2-based RDTs remain positive for several weeks after parasite clearance whereas microscopy detects only circulating parasites, we refitted the model under two alternative malaria definitions, positivity on both assays, and microscopy positivity alone. Agreement between the two assays was summarised by Cohen’s *κ*. All three specifications used identical marginal equations and differed only in the definition of *Y_M_* . Finally, we assessed robustness of the significant marginal associations to unmeasured con-founding by computing E-values with the EValue R package [65, 66]. Methods for the analyses of infection intensity, haemoglobin and anaemia are given in Supplementary Note 3.

## Data availability

The raw data are protected and are not available due to data privacy laws. De-identified data can be requested from the corresponding author (G.F.C.) and will be made available, subject to approval by the relevant ethics committees and a data-sharing agreement. Sentinel-2 imagery is publicly available from the Copernicus Data Space Ecosystem [46].

## Code availability

The R code has been provided as supplementary material.

## Data Availability

All data produced in the present study are available upon reasonable request to the authors

## Acknowledgements

We are grateful for the engagement and participation of all study participants and local communities involved in this research. Without our field teams this research would not be possible, and we extend our thanks to everyone involved. We thank the SchistoTrack group for valuable feedback and insights during group meetings and conversations.

## Author contributions

Conceptualisation: M.M.L., C.A.D. and G.F.C. Data curation: M.M.L., V.T., E.A., R.B., A.M., B.I., J.D.B., B.N., N.B.K. and G.F.C. Formal analysis: M.M.L. Funding acquisition: M.M.L. and G.F.C. Investigation: M.M.L. Methodology: M.M.L., C.A.D. and G.F.C. Project administration: N.B.K. and G.F.C. Resources: N.B.K. and G.F.C. Software: M.M.L., G.F.C. Supervision: C.A.D. and G.F.C. Validation: M.M.L. Visualisation: M.M.L. Writing (original draft): M.M.L. Writing (review and editing): M.M.L., V.T., E.A., R.B., A.M., B.I., J.D.B., B.N., N.B.K., C.A.D. and G.F.C. All authors read and approved the final manuscript.

## Ethics declarations

### Ethics approval

After review data collection and use were approved by the Oxford Tropical Research Ethics Committee (OxTREC 509-21), Vector Control Division Research Ethics Committee of the Uganda Ministry of Health (VCDREC146), and Uganda National Council of Science and Technology (UNCST HS 1664ES). All adult participants provided written informed consent. For children, informed assent was obtained from the child and written informed consent from a parent or legal guardian.

