## Supplementary Information for "Identifying shared and parasite-specific determinants of *Plasmodium falciparum* and *Schistosoma mansoni* co-infection in a large-scale community-based study in rural Uganda"

### Supplementary Notes

#### Supplementary Note 1: Copula model specification and estimands

##### Copulas for binary random variables

For the following, consider Bernoulli variables $Y_1, Y_2 \in \{0, 1\}$ with marginal probabilities $p_k = P(Y_k = 1) \in (0, 1)$, $k = 1, 2$, and cell probabilities $p_{ij} = P(Y_1 = i, Y_2 = j)$. The CDF of both margins takes only three values, $\mathrm{Ran}(F_{Y_k}) = \{0,\ 1 - p_k,\ 1\}$, so the grid $\mathrm{Ran}(F_{Y_1}) \times \mathrm{Ran}(F_{Y_2})$ contains a single interior constraint, which is $C(1 - p_1, 1 - p_2) = p_{00}$. A $2 \times 2$ table can be reproduced by any copula family that spans the full Fréchet range, such that the family cannot be chosen from binary data and must be justified on structural grounds [1].

##### Plackett copula

Plackett [2] defined a family of bivariate distributions by requiring that the cross-product ratio of the $2 \times 2$ table formed by dichotomising both margins at any point $(u, v) \in (0, 1)^2$ equals a constant $\theta > 0$:

$$\theta = \frac{C(u, v)\,[1 - u - v + C(u, v)]}{[u - C(u, v)]\,[v - C(u, v)]}, \tag{S1}$$

then writing $t = C(u, v)$ rearranges this to

$$(\theta - 1)\,t^2 - [1 + (\theta - 1)(u + v)]\,t + \theta\,uv = 0, \tag{S2}$$

within the admissible range the root gives

$$C(u, v; \theta) = \frac{[1 + (\theta - 1)(u + v)] - \sqrt{[1 + (\theta - 1)(u + v)]^2 - 4uv\,\theta(\theta - 1)}}{2(\theta - 1)} \tag{S3}$$

for $\theta \neq 1$. One can easily show that in case $\theta = 1$ , $t = uv$, recovering the independence copula $\Pi(u, v) = uv$.

Evaluating (S1) at the Sklar constraint point $(u, v) = (1 - p_1, 1 - p_2)$, where $C(u, v) = p_{00}$, the four cells reduce to $p_{00}$, $p_{01}$, $p_{10}$, $p_{11}$, and the equation becomes $\theta = p_{00}\, p_{11}/(p_{01}\, p_{10}) = \psi$. The Plackett copula parameter is therefore algebraically identical to the cross-product OR [2, 1].

#### Cell probabilities and the joint AME

Given the marginal probabilities $p_S$ and $p_M$ and copula parameter $\psi$, the four cell probabilities are:

$$\begin{aligned} p_{11} &= C_\psi(p_S, p_M), \\ p_{10} &= p_S - p_{11}, \\ p_{01} &= p_M - p_{11}, \\ p_{00} &= 1 - p_S - p_M + p_{11}. \end{aligned}$$

The co-infection AME is obtained as:

$$\widehat{\text{AME}}_{11}(Z) = \frac{1}{N} \sum_{i=1}^{N} \big[C_{\hat\psi}(\hat p_{S,i}^{(1)}, \hat p_{M,i}^{(1)}) - C_{\hat\psi}(\hat p_{S,i}^{(0)}, \hat p_{M,i}^{(0)})\big]. \tag{S4}$$

#### Estimands and G-computation

For each individual $i$ the fitted copula model yields risk $\hat p_{S,i}$, $\hat p_{M,i}$, and the joint co-infection risk $\hat p_{11,i} = C_{\hat\psi}(\hat p_{S,i}, \hat p_{M,i})$. When $\hat\psi > 1$, the copula adjusts the joint probability upward relative to the independence product $\hat p_{S,i} \hat p_{M,i}$ and in case $\hat\psi < 1$, the joint probability is adjusted downward.

#### Conditional AMEs

In our setting we are primarily interested in two quantities, the predicted risk attributable to the exposure for those who currently report it and secondly the predicted risk reduction if an intervention were extended to those who currently lack the exposure or protective factor. The latter is the more directly relevant quantity for programme rollout. These are the observational analogues of the average treatment effect on the treated and untreated [3], however, although we do adopt that terminology, we note that exposures here are self-selected, not randomised.

The overall AME is the prevalence-weighted average of the two conditional AMEs, which for binary $Z$ with sample prevalence $\pi$ is $\text{AME} = \pi \cdot \text{AME}|_{Z=1} + (1 - \pi) \cdot \text{AME}|_{Z=0}$.

#### Population attributable fraction

The PAF for a binary covariate $Z$ is

$$\widehat{\text{PAF}}_d(Z) = \frac{\bar{p}_d^{\text{obs}} - \bar{p}_d^{(Z=0)}}{\bar{p}_d^{\text{obs}}}, \tag{S5}$$

where $\bar{p}_d^{\text{obs}} = N^{-1} \sum_i \hat{p}_{d,i}$ and $\bar{p}_d^{(Z=0)} = N^{-1} \sum_i \hat{p}_{d,i}^{(0)}$.

Given that interventions are typically delivered as packages, we also considered a joint PAF, which sets all covariates in a block to their target levels and computes equation S5 from the resulting scenario predictions. Formally, for a block $\mathcal{B} = \{Z_1, \dots, Z_K\}$ under action $a$ this gives:

$$\widehat{\text{PAF}}_d(\mathcal{B}, a) = \frac{\bar{p}_d^{\text{obs}} - \bar{p}_d^{(\mathcal{B}=a)}}{\bar{p}_d^{\text{obs}}}. \tag{S6}$$

We hold all other covariates at the observed values, such that $\bar{p}_d^{(\mathcal{B}=a)}$ is the average predicted probability when every variable in $\mathcal{B}$ is set as the action specified.

### Supplementary Note 2: Satellite shoreline detection pipeline

#### Hexagonal grid framework

For our satellite imagery analysis we followed a similar approach as in Lang et al. [4]. We used a tessellated study area based on a hexagonal cell grid of approximately 900 m$^2$.

For each hexagon $h$, the MNDWI was computed from median composites of cloud-free Sentinel-2 Level-2A scenes (December–January periods, 2021–2024):

$$\text{MNDWI}_h = \frac{\text{Green}_h - \text{SWIR}_h}{\text{Green}_h + \text{SWIR}_h}, \tag{S7}$$

where Green is Band 3 and SWIR is Band 11 (short-wave infrared), and the per-pixel index was averaged within each hexagon. Values above zero indicate likely water and below zero indicate land.

#### Water probability

Each hexagon was assigned a local water probability by a sigmoid transformation of its MNDWI:

$$\phi_h^{(0)} = \frac{1}{1 + \exp\bigl(-k\,\mathrm{MNDWI}_h\bigr)}. \tag{S8}$$

The steepness of the sigmoid function was set to $k = 10$ and the function was additionally centred on MNDWI $= 0$, which is the conventional water–land boundary of the index.

#### Classification and shoreline extraction

After we obtained the probabilities from the sigmoid function, we thresholded them by Otsu's method [5], which selects the value minimising the within-class variance:

$$\tau^* = \arg\min_{\tau}\bigl[\omega_0(\tau)\,\sigma_0^2(\tau) + \omega_1(\tau)\,\sigma_1^2(\tau)\bigr]. \tag{S9}$$

Here, $\omega_0(\tau)$ and $\omega_1(\tau)$ are the proportions of hexagons with smoothed probability below and above $\tau$, and $\sigma_0^2(\tau)$ and $\sigma_1^2(\tau)$ are the corresponding within-class variances. We also evaluated the threshold over a 256-bin histogram of $\phi$, equivalently by maximising the between-class variance $\omega_0\omega_1(\mu_0 - \mu_1)^2$. Each hexagon was classified as water if $\phi_h > \tau^*$ and as land otherwise, giving $w_h \in \{0, 1\}$.

Shoreline hexagons were then identified topologically as those lying on a water–land boundary.

$$S_h = \nVdash\Bigl\{\,\bigl|\,w_h - \overline{w_{N(h)}}\,\bigr| > 0\,\Bigr\}. \tag{S10}$$

Any hexagon having at least one adjacent hexagon of the opposite class (water or land) is automatically classified as shoreline. Distance from each household to the nearest shoreline hexagon was computed as the Euclidean distance between the household coordinates and the nearest shoreline hex centroid, and entered the schistosomiasis margin as $\log(1 + d)$ with $d$ in metres, on the same transform as the field-mapped distance.

### Supplementary Note 3: Infection intensity, haemoglobin and anaemia analyses

#### Infection intensity

Descriptive intensity analyses were nested within the primary study population. For *P. falciparum* measurable parasitaemia was required ($> 0$ parasites/$\mu$L), and for *S. mansoni* intensity a measurable egg count ($> 0$ EPG). We compared geometric mean EPG and geometric mean parasitaemia between mono- and co-infected individuals by Welch's $t$-test on the log-transformed intensity. Age profiles of intensity were estimated with a penalised thin-plate spline of age on log intensity, back-transformed to the original scale, with pointwise 95% CIs from a village-level bootstrap. The results are shown in Supplementary Figs. S9, S10 and S11.

#### Interaction on haemoglobin and anaemia

We tested whether the effect of co-infection on haemoglobin was the sum of the two single-infection effects by regressing altitude-adjusted haemoglobin on indicators of EPG-positive (*S. mansoni*) and *P. falciparum* infection and their interaction. The model was fitted unadjusted and adjusted for age, gender and village, with age as a penalised thin-plate spline and village as a random effect [6]. For any anaemia, we summarised deviation from additivity by the relative excess risk due to interaction ($\text{RERI} = \text{RR}_{11} - \text{RR}_{10} - \text{RR}_{01} + 1$) [7], with risk ratios from a log-Poisson model with the same exposures and adjustment. Additionally PAFs quantified the proportional reduction in anaemia prevalence if all participants were uninfected with *S. mansoni*, *P. falciparum* or both, with all other covariates held at their observed values. CIs for the RERI and PAFs were obtained by nonparametric bootstrap ($B = 5000$), resampling villages in the adjusted analyses. Results are shown in Supplementary Fig. S12 and Supplementary Table S11. Moreover, we obtained soil-transmitted helminths (STH) from the same Kato-Katz slides and refitted the adjusted haemoglobin, RERI and PAF models in Mayuge with any STH or hookworm infection (egg count $> 0$) as a covariate. The analysis was limited to Mayuge because STH infection was common only there (Supplementary Table S12).

#### Haemoglobin measurement and anaemia classification

Capillary haemoglobin (Hb, g/dL) was measured at the clinical stations from the same finger-prick sample used for malaria RDT. We encoded any value outside the biologically plausible range of 4–22 g/dL to missing.

Measured Hb was adjusted for elevation prior to anaemia classification, using the continuous adjustment recommended by the WHO 2024 guideline on haemoglobin cutoffs [8]:

$$\text{Hb}_{\text{adj}} = \text{Hb}_{\text{meas}} - \frac{0.0056384\,E + 0.0000003\,E^2}{10}, \tag{S11}$$

where $E$ is elevation in metres and division by 10 converts the adjustment from g/L to g/dL. Because single-household GPS altitude readings were noisy, each participant was assigned the median GPS altitude of the study households in their village. Village elevations ranged from 1141 to 1185 m in Mayuge, 619 to 635 m in Buliisa and 623 to 661 m in Pakwach. Of the 4278 participants entering the anaemia analysis, 1070 (25.01%), all in Mayuge, lived above 1000 m. The mean correction across the sample was −0.45 g/dL and the largest was −0.71 g/dL.

Anaemia was classified from altitude-adjusted Hb using the WHO 2024 thresholds [8]. For participants aged 5 years and older these thresholds are unchanged from the WHO 2011 reference [9]. The 2024 update revised only the under-5 cutoffs, which do not apply to this cohort (minimum age 5 years). Age-, gender-, and pregnancy-specific cutoffs for normal, mild, moderate, and severe anaemia are in Supplementary Table S10. Pregnancy status was taken from the sonography survey [10].

We excluded nine participants with missing haemoglobin, and three women aged 15 years or older whose pregnancy status was unknown and who could therefore not be assigned a WHO anaemia category, since the cutoffs differ for pregnant and non-pregnant women. This yielded $n = 4278$, 12 fewer than the main analytic sample. Mean Hb was compared across infection groups using Welch's $t$-test on the altitude-adjusted scale [11]. The prevalence of any anaemia (mild, moderate, or severe combined) was compared across the four infection groups by Pearson's $\chi^2$ test on the resulting $4 \times 2$ table.

### Supplementary Tables

**Table S1**: **Snails and sanitation at mapped water-contact sites by water landform.** Sanitation percentages are of sites within each landform, and shedding percentages are of snails collected within each landform. Rows are ordered by the percentage of snails shedding cercariae.

| Water landform | Sites ($n$) | Sanitation activity $n$ (%) | Snails collected | Snails per site | Snails shedding $n$ (%) |
|---|---|---|---|---|---|
| Lake beach | 166 | 88 (53.0) | 15,575 | 93.8 | 228 (1.46) |
| River or river marsh | 69 | 22 (31.9) | 5,249 | 76.1 | 43 (0.82) |
| Lake marsh | 189 | 65 (34.4) | 18,843 | 99.7 | 102 (0.54) |
| Pond or swamp | 65 | 13 (20.0) | 6,401 | 98.5 | 10 (0.16) |
| Rice paddy | 10 | 2 (20.0) | 2,094 | 209.4 | 1 (0.05) |
| Total | 499 | 190 (38.1) | 48,162 | 96.5 | 384 (0.80) |

**Table S2**: **Models allowing residual dependence to vary.** Each model adds a linear predictor for $\log \psi_i$ to the constant-$\psi$ baseline (M0), with both model margins unchanged. ΔAIC is relative to M0, and positive values indicate worse fit. The last columns summarise fitted $\hat{\psi}_i$ across participants. $s(\cdot)$, penalised spline; adult, aged $\geq 18$ years; water distance, $\ln(1 + \text{metres})$ to the nearest water-contact site; EDF, effective degrees of freedom; $\log L$, log-likelihood.

| | | | | | | $\hat{\psi}_i$ | |
|---|---|---|---|---|---|---|---|
| Model | Predictor for $\log \psi_i$ | EDF | $\log L$ | AIC | ΔAIC | Median | Range |
| M0 | Constant | 150.6 | −4785.80 | 9872.7 | – | 1.183 | – |
| M1a | $s$(age) | 155.2 | −4782.10 | 9874.6 | 1.86 | 1.203 | 0.210–1.595 |
| M1b | District | 152.6 | −4785.64 | 9876.5 | 3.73 | 1.198 | 1.090–1.229 |
| M1c | Gender | 151.6 | −4785.71 | 9874.6 | 1.87 | 1.155 | 1.155–1.212 |
| M2 | $s$(age) + district | 157.2 | −4782.04 | 9878.4 | 5.70 | 1.200 | 0.201–1.631 |
| M3 | $s$(age) + district + gender | 158.2 | −4781.94 | 9880.3 | 7.58 | 1.197 | 0.196–1.678 |
| M4a | $s$(age) by district + district | 155.7 | −4783.81 | 9879.0 | 6.26 | 1.199 | 0.443–2.749 |
| M4b | M3 + adult × gender | 160.9 | −4778.95 | 9879.7 | 6.97 | 1.149 | 0.129–2.229 |
| M5a | $s$(water distance) | 151.7 | −4785.62 | 9874.6 | 1.91 | 1.193 | 1.089–1.216 |
| M5b | $s$(water distance) + district | 154.0 | −4785.16 | 9878.2 | 5.50 | 1.207 | 0.965–1.270 |

**Table S3**: **Residual dependence by district.** The bivariate model was refitted separately within each district. CIs are back-transformed Wald intervals on $\log \hat{\psi}$. Estimates were homogeneous across districts (Cochran's $Q = 0.69$, df = 2, $p = 0.71$), and no pairwise contrast differed ($p \geq 0.41$).

| District | $n$ | $\log \hat{\psi}$ (SE) | $\hat{\psi}$ (95% CI) | $\Upsilon$ |
|---|---|---|---|---|
| Mayuge | 1074 | 0.041 (0.181) | 1.04 (0.73, 1.48) | 0.01 |
| Buliisa | 1457 | 0.234 (0.147) | 1.26 (0.95, 1.69) | 0.06 |
| Pakwach | 1759 | 0.167 (0.124) | 1.18 (0.93, 1.51) | 0.04 |
| Pooled (main model) | 4290 | 0.168 (0.081) | 1.18 (1.01, 1.39) | 0.04 |

**Table S4**: **Backward elimination of candidate covariates.** Covariates are listed in the order they were removed from the bivariate model. The candidate set included the home quality score, home ownership, room count, household size, majority religion, majority tribe, household social status, log-distance to nearest tap or latrine, water landform type and log-distance to nearest water-contact site. These entered both margins and were subject to backward elimination. All other covariates were retained in every model (Methods).

| Step | Covariate dropped | AIC before | AIC after | ΔAIC |
|---|---|---|---|---|
| 1 | Home owned | 9885.4 | 9881.6 | −3.81 |
| 2 | Home quality score | 9881.6 | 9878.4 | −3.16 |
| 3 | Household social status | 9878.4 | 9876.3 | −2.15 |
| 4 | Household size | 9876.3 | 9874.3 | −2.02 |
| 5 | Distance to nearest tap or latrine | 9874.3 | 9872.7 | −1.55 |
| Final model | | | 9872.7 | |

**Table S5**: **Sensitivity to household clustering.** With one adult and one child per household, the design effect is $1 + \rho$, where $\rho$ is the household intraclass correlation (ICC). (**a**) Variance components from intercept-only logistic models with village and household random intercepts. ICCs are on the latent scale, and the household variance for *P. falciparum* was estimated at zero. (**b**) The bivariate model refitted in subsamples with one individual per household. The two estimates were homogeneous (Cochran's $Q = 0.02$, df $= 1$, $p = 0.88$).

**a** Variance components and intraclass correlations

| | Random-intercept variance | | ICC | | |
|---|---|---|---|---|---|
| | Household | Village | Household | Village | Design effect |
| *S. mansoni* | 0.149 | 0.396 | 0.039 | 0.103 | 1.04 |
| *P. falciparum* | 0.000 | 0.360 | 0.000 | 0.099 | 1.00 |

**b** Refits in samples with one individual per household

| | $n$ | Individuals per household | $\hat{\psi}$ (95% CI) |
|---|---|---|---|
| All participants (main model) | 4290 | 2 | 1.18 (1.01, 1.39) |
| Adults only | 2149 | 1 | 1.23 (0.97, 1.56) |
| Children only | 2141 | 1 | 1.20 (0.96, 1.51) |

**Table S6**: **Agreement between malaria RDT and blood-film microscopy** ($n = 4290$). Parasite density categories follow WHO definitions. Agreement statistics use the 4201 participants with both results. Observed agreement was 80.05% (Cohen's $\kappa = 0.58$). With microscopy as reference, RDT sensitivity was 82.57%, specificity 78.83%, positive predictive value 65.40% and negative predictive value 90.32%. RDT-positive, microscopy-negative results are consistent with HRP2 antigen persisting after parasite clearance.

| Microscopy | RDT negative | RDT positive | Total | RDT positive (%) |
|---|---|---|---|---|
| Negative (no parasites detected) | 2231 | 599 | 2830 | 21.2 |
| Positive | 239 | 1132 | 1371 | 82.6 |
| Low density | 222 | 760 | 982 | 77.4 |
| Moderate density | 12 | 248 | 260 | 95.4 |
| High density | 5 | 124 | 129 | 96.1 |
| Both results available | 2470 | 1731 | 4201 | 41.2 |
| No microscopy result | 53 | 36 | 89 | 40.4 |
| Total | 2523 | 1767 | 4290 | 41.2 |

**Table S7**: **Residual dependence under three malaria definitions.** The bivariate model was refitted with identical marginal equations, changing only the malaria outcome. Sample sizes differ because RDT-negative participants are negative under the combined definition regardless of microscopy. CIs are Wald intervals on $\log \hat{\psi}$. Agreement between the two assays is shown in Supplementary Table S6.

| Malaria definition | $n$ | Positive, $n$ (%) | $\hat{\psi}$ (95% CI) | $\Upsilon$ |
|---|---|---|---|---|
| RDT positive (primary) | 4290 | 1767 (41.2) | 1.18 (1.01, 1.39) | 0.04 |
| RDT positive and microscopy positive | 4290 | 1132 (26.4) | 1.06 (0.89, 1.26) | 0.02 |
| Microscopy positive | 4201 | 1371 (32.6) | 1.03 (0.88, 1.21) | 0.01 |

**Table S8**: **Performance of the main and shoreline models.** The shoreline model replaces distance to the nearest field-mapped water-contact site with distance to the satellite-derived shoreline and omits water landform.

| | Field-mapped (main model) | | | Satellite-derived shoreline | | |
|---|---|---|---|---|---|---|
| | *S. mansoni* | *P. falciparum* | Co-infection | *S. mansoni* | *P. falciparum* | Co-infection |
| *Discrimination* | | | | | | |
| AUC, pooled | 0.712 | 0.786 | 0.759 | 0.713 | 0.785 | 0.759 |
| AUC, fold mean | 0.712 | 0.786 | 0.760 | 0.713 | 0.785 | 0.760 |
| AUC, fold SD | 0.016 | 0.027 | 0.021 | 0.016 | 0.026 | 0.022 |
| *Calibration* | | | | | | |
| Brier score | 0.210 | 0.183 | 0.129 | 0.210 | 0.184 | 0.130 |
| Intercept | −0.027 | −0.012 | −0.065 | −0.025 | −0.013 | −0.076 |
| Slope | 0.914 | 0.952 | 0.944 | 0.922 | 0.953 | 0.940 |
| *Classification* | | | | | | |
| Sensitivity | 0.655 | 0.726 | 0.666 | 0.668 | 0.686 | 0.679 |
| Specificity | 0.655 | 0.707 | 0.711 | 0.633 | 0.746 | 0.699 |
| Balanced accuracy | 0.655 | 0.716 | 0.688 | 0.651 | 0.716 | 0.689 |
| Positive predictive value | 0.574 | 0.634 | 0.343 | 0.563 | 0.654 | 0.338 |
| Negative predictive value | 0.728 | 0.787 | 0.904 | 0.729 | 0.773 | 0.905 |
| *Full-data fit* | | | | | | |
| AIC | | 9872.7 | | | 9881.3 ($\Delta = +8.6$) | |
| $\hat{\psi}$ (95% CI) | | 1.18 (1.01, 1.39) | | | 1.18 (1.01, 1.38) | |

**Table S9: E-values for associations with 95% CIs excluding 1.** The E-value is the minimum risk ratio an unmeasured confounder would need with both exposure and outcome to explain away the observed OR. The CI E-value applies to the limit closest to 1. ORs below 1 were inverted, and the square-root approximation for common outcomes was used. Distances are per unit of $\ln(1 + \text{metres})$.

| | | | E-value | |
|---|---|---|---|---|
| | OR (95% CI) | $p$ | Estimate | CI |
| ***S. mansoni*** | | | | |
| Female (ref: male) | 0.85 (0.72, 1.00) | 0.045 | 1.39 | 1.05 |
| Years of education | 0.96 (0.93, 0.99) | 0.009 | 1.17 | 1.08 |
| Village majority religion | 1.27 (1.07, 1.51) | 0.006 | 1.51 | 1.23 |
| Village majority tribe | 1.27 (1.06, 1.52) | 0.011 | 1.50 | 1.20 |
| Fishing in open water | 1.63 (1.25, 2.14) | <0.001 | 1.88 | 1.48 |
| Treats drinking water | 0.80 (0.67, 0.96) | 0.018 | 1.48 | 1.16 |
| Distance to health centre | 1.32 (1.12, 1.56) | <0.001 | 1.57 | 1.31 |
| Distance to water-contact site | 0.80 (0.71, 0.91) | <0.001 | 1.48 | 1.28 |
| Pond or swamp (ref: lake beach) | 0.65 (0.48, 0.88) | 0.005 | 1.78 | 1.34 |
| River or river marsh (ref: lake beach) | 0.67 (0.48, 0.94) | 0.021 | 1.74 | 1.21 |
| Recruitment year 2023 (ref: 2022) | 0.64 (0.44, 0.92) | 0.016 | 1.82 | 1.25 |
| Recruitment year 2024 (ref: 2022) | 1.56 (1.19, 2.05) | 0.001 | 1.81 | 1.40 |
| ***P. falciparum*** | | | | |
| Female (ref: male) | 0.76 (0.64, 0.90) | 0.001 | 1.56 | 1.30 |
| Farmer (ref: none/other) | 0.76 (0.59, 0.98) | 0.033 | 1.56 | 1.12 |
| Distance to drug shop | 1.11 (1.01, 1.22) | 0.037 | 1.29 | 1.06 |
| Distance to health centre | 1.34 (1.15, 1.57) | <0.001 | 1.59 | 1.35 |
| NDVI (vegetation) | 2.50 (1.00, 6.22) | 0.050 | 2.54 | 1.03 |
| Lake marsh (ref: lake beach) | 1.61 (1.24, 2.10) | <0.001 | 1.85 | 1.47 |
| Pond or swamp (ref: lake beach) | 1.78 (1.31, 2.41) | <0.001 | 2.00 | 1.55 |
| River or river marsh (ref: lake beach) | 1.81 (1.28, 2.56) | <0.001 | 2.03 | 1.51 |
| Recruitment year 2024 (ref: 2022) | 0.30 (0.21, 0.41) | <0.001 | 3.07 | 2.49 |

**Table S10: WHO 2024 haemoglobin cutoffs (g/dL) for anaemia in individuals aged 5 years and over.** Altitude-adjusted haemoglobin was compared against these cutoffs, which are unchanged from the 2011 reference for these age groups.

| Group | Severe | Moderate | Mild | Normal |
|---|---|---|---|---|
| Children 5–11 y | $< 8.0$ | 8.0–10.9 | 11.0–11.4 | $\geq 11.5$ |
| Children 12–14 y | $< 8.0$ | 8.0–10.9 | 11.0–11.9 | $\geq 12.0$ |
| Non-pregnant women 15+ y | $< 8.0$ | 8.0–10.9 | 11.0–11.9 | $\geq 12.0$ |
| Pregnant women 15+ y | $< 7.0$ | 7.0–9.9 | 10.0–10.9 | $\geq 11.0$ |
| Men 15+ y | $< 8.0$ | 8.0–10.9 | 11.0–12.9 | $\geq 13.0$ |

**Table S11**: **Interaction between infections on haemoglobin and anaemia** ($n = 4278$). Adjusted models include age (penalised spline), gender and a village random effect. Haemoglobin differences are in g/dL with Wald 95% CIs. CIs for RERI and PAF are from a village-level bootstrap. Anaemia follows WHO 2024 cutoffs. PAF, population attributable fraction; RERI, relative excess risk due to interaction.

| | Crude<br>Estimate (95% CI) | Adjusted<br>Estimate (95% CI) |
|---|---|---|
| *Haemoglobin (g/dL)* | | |
| *S. mansoni* | −0.04 (−0.21, 0.13) | −0.07 (−0.23, 0.09) |
| *P. falciparum* | −0.95 (−1.12, −0.77) | −0.33 (−0.50, −0.17) |
| *S. mansoni* × *P. falciparum* | 0.15 (−0.11, 0.41) | 0.04 (−0.20, 0.28) |
| *Additive interaction, any anaemia* | | |
| RERI | −0.08 (−0.20, 0.02) | −0.05 (−0.16, 0.04) |
| *PAF of any anaemia (%)* | | |
| All infections | 15.5 (11.7, 18.7) | 8.8 (5.6, 12.4) |
| *S. mansoni* only | 1.2 (−0.6, 2.9) | 0.8 (−1.0, 2.5) |
| *P. falciparum* only | 8.0 (6.3, 10.0) | 4.5 (2.8, 6.5) |
| Co-infection | 6.3 (4.4, 8.0) | 3.5 (2.1, 5.0) |

**Table S12**: **Haemoglobin and anaemia adjusted for soil-transmitted helminths (STH) in Mayuge** ($n = 1070$). STH prevalence by Kato-Katz was 18.0% in Mayuge (hookworm 17.2%), 2.4% in Buliisa and 0.9% in Pakwach. All models adjust for age, gender and village, and STH-adjusted models add any STH or hookworm infection. CIs are Wald (haemoglobin) or delta-method (RERI) intervals.

| | Not adjusted<br>for STH | + any<br>STH | +<br>hookworm |
|---|---|---|---|
| *Haemoglobin difference, g/dL (95% CI)* | | | |
| *S. mansoni* | 0.18<br>(−0.08 to 0.44) | 0.18<br>(−0.08 to 0.44) | 0.18<br>(−0.08 to 0.44) |
| *P. falciparum* | −0.25<br>(−0.53 to 0.02) | −0.25<br>(−0.53 to 0.02) | −0.25<br>(−0.53 to 0.02) |
| Interaction | −0.20<br>(−0.62 to 0.23) | −0.20<br>(−0.62 to 0.23) | −0.20<br>(−0.62 to 0.23) |
| *Any anaemia* | | | |
| RERI (95% CI) | 0.04<br>(−0.34 to 0.41) | 0.04<br>(−0.34 to 0.41) | 0.04<br>(−0.34 to 0.41) |
| PAF, all infections (%) | 5.94 | 5.92 | 5.89 |
| PAF, *S. mansoni* only (%) | −0.84 | −0.85 | −0.86 |
| PAF, *P. falciparum* only (%) | 4.36 | 4.35 | 4.34 |
| PAF, co-infection (%) | 2.42 | 2.41 | 2.40 |

### Supplementary Figures

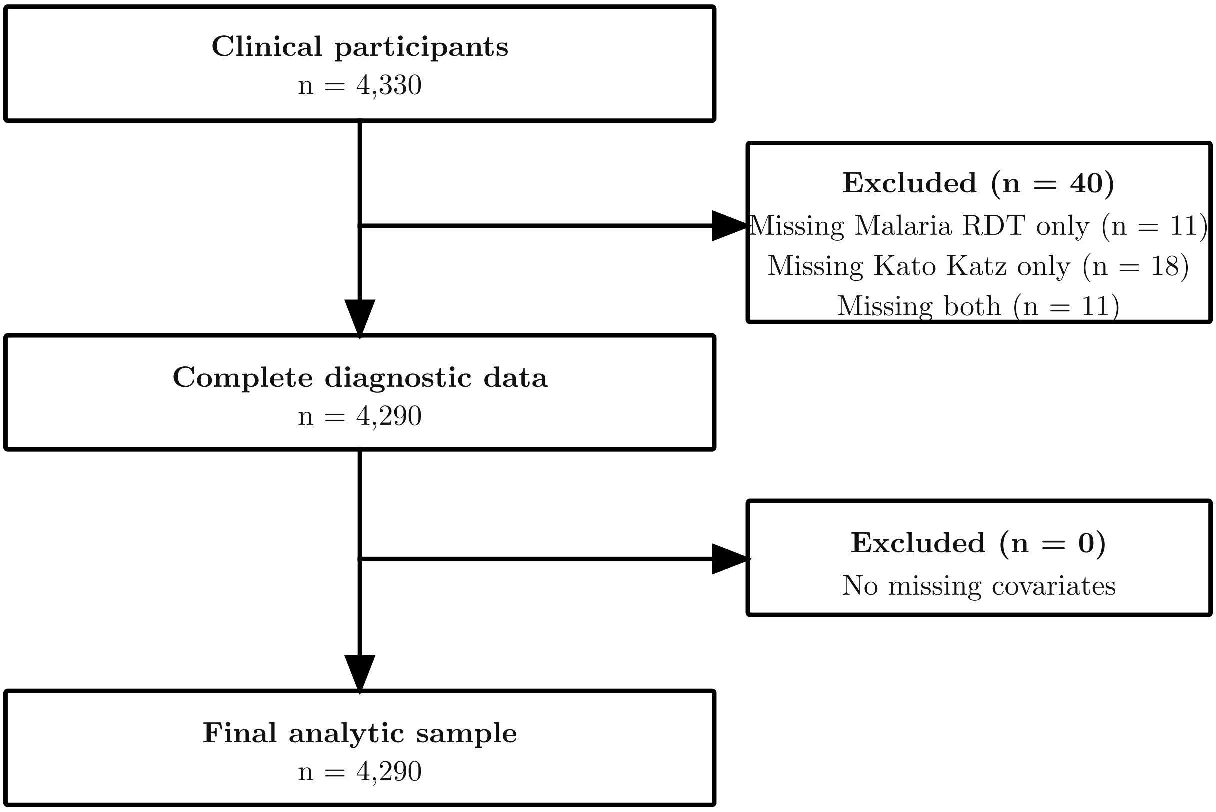


**Fig. S1**: **Flowchart of participants.** Of 4330 participants enrolled in SchistoTrack between 2022 and 2024, 40 were excluded for missing diagnostic results (11 missing RDT, 18 missing Kato-Katz and 11 missing both), leaving 4290 for analysis.

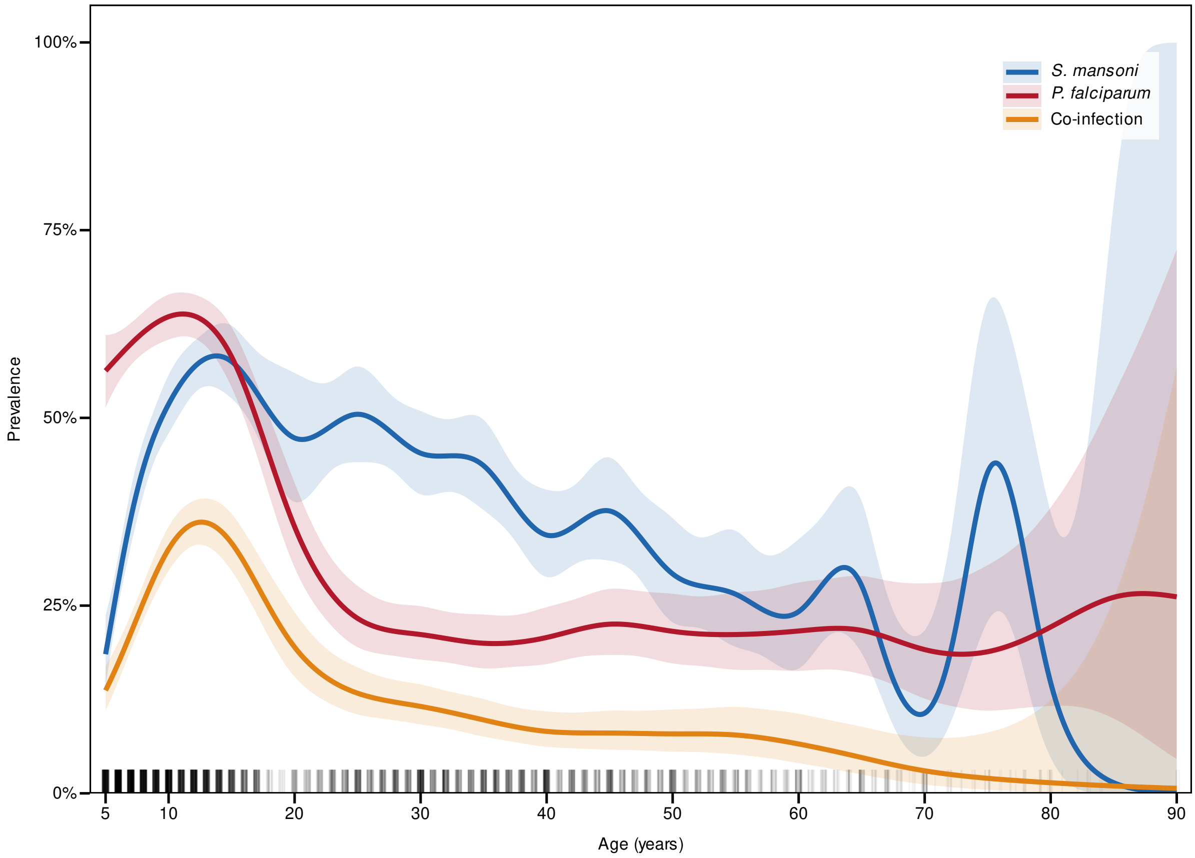


**Fig. S2**: **Unadjusted age-prevalence curves** for *S. mansoni*, *P. falciparum* and co-infection, from generalised additive models with 95% CIs.

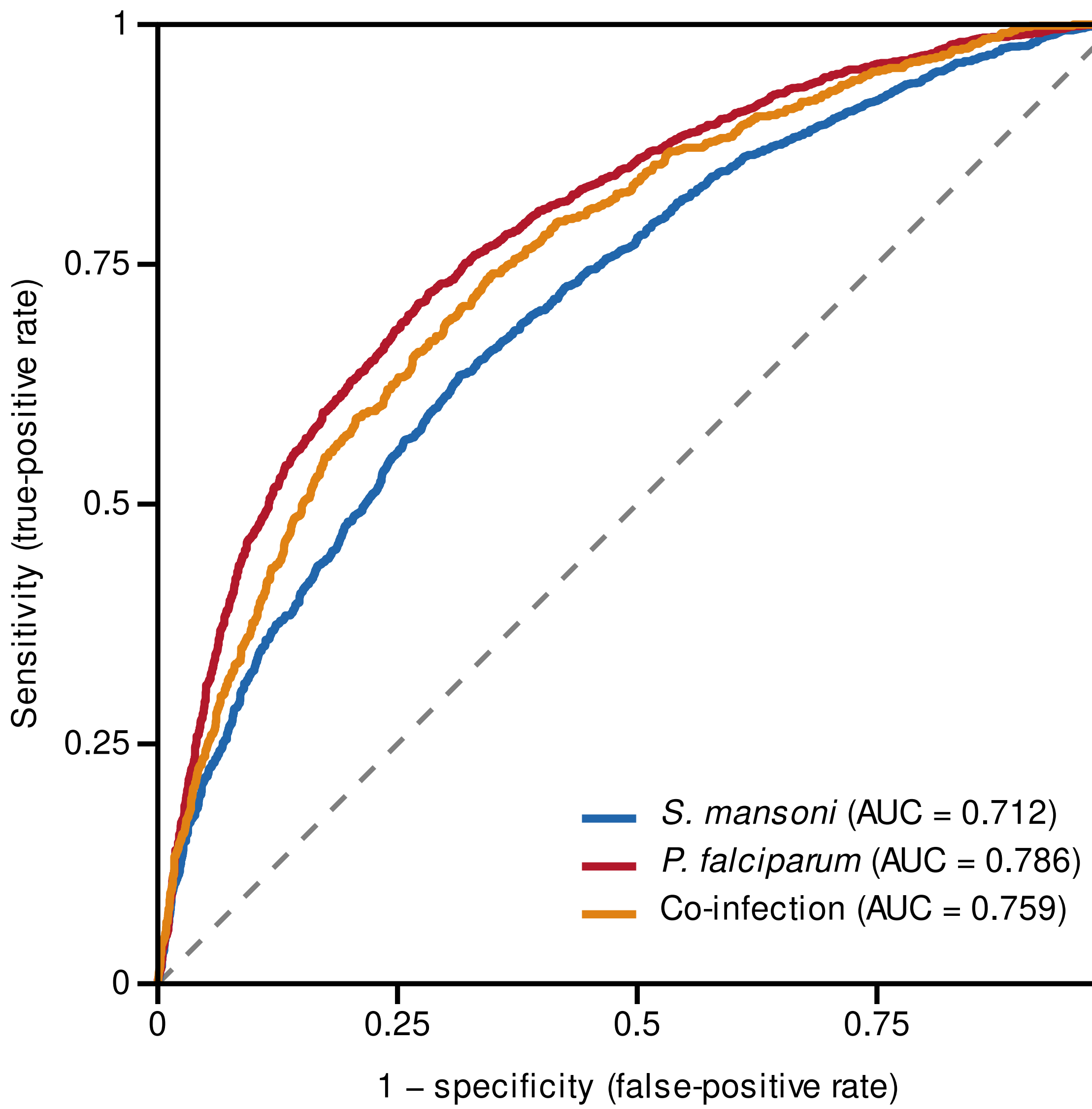


**Fig. S3**: **Receiver operating characteristic curves for the main model under 10-fold cross-validation.** Predictions were pooled across held-out folds. The area under the curve (AUC) was 0.712 for *S. mansoni*, 0.786 for *P. falciparum* and 0.759 for co-infection.

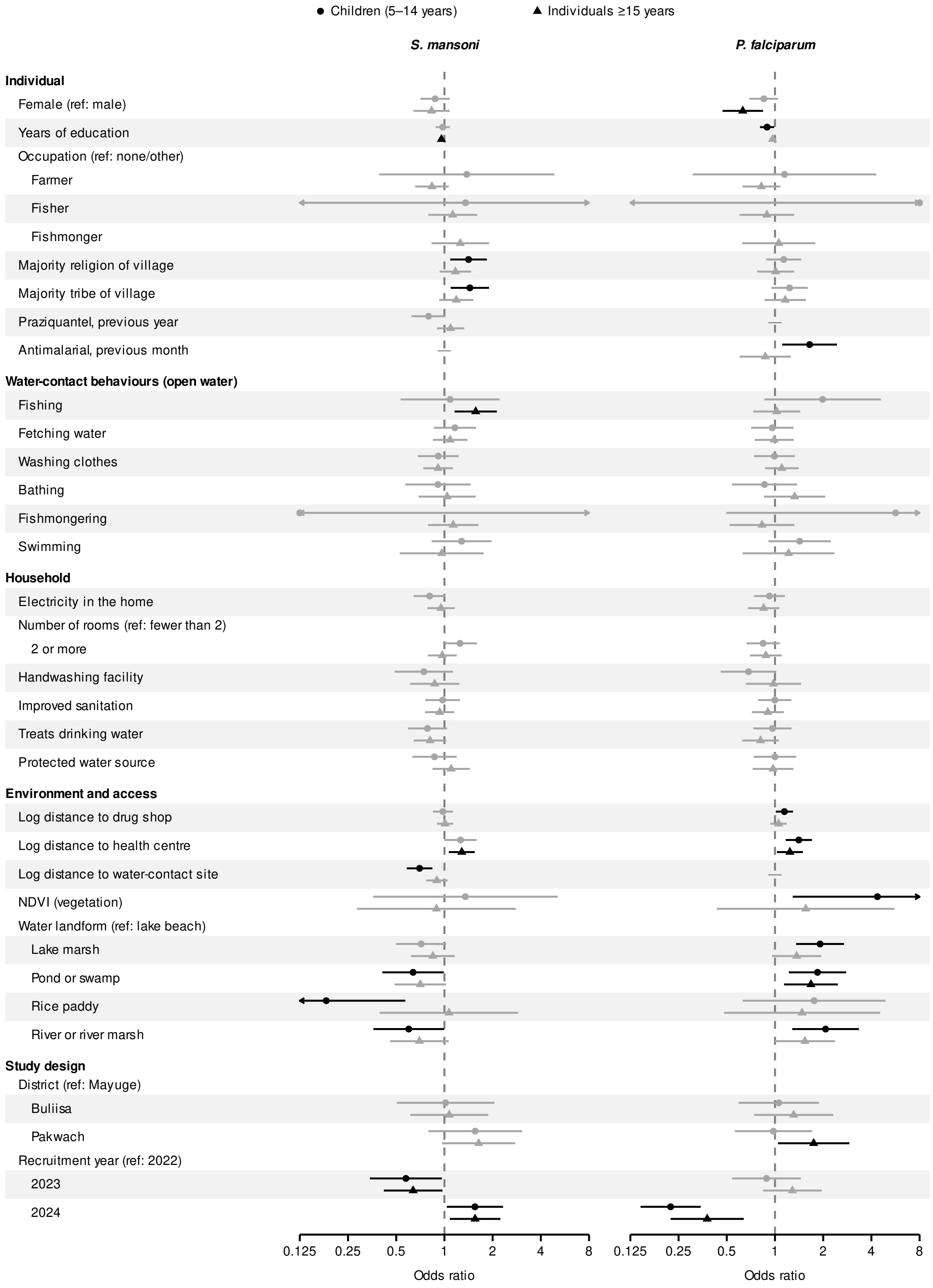


**Fig. S4**: **Adjusted odds ratios by age group.** Bivariate copula models fitted separately to children aged 5–14 years ($n = 1905$, circles) and individuals aged 15 years or older ($n = 2385$, triangles). Black points have $p < 0.05$.

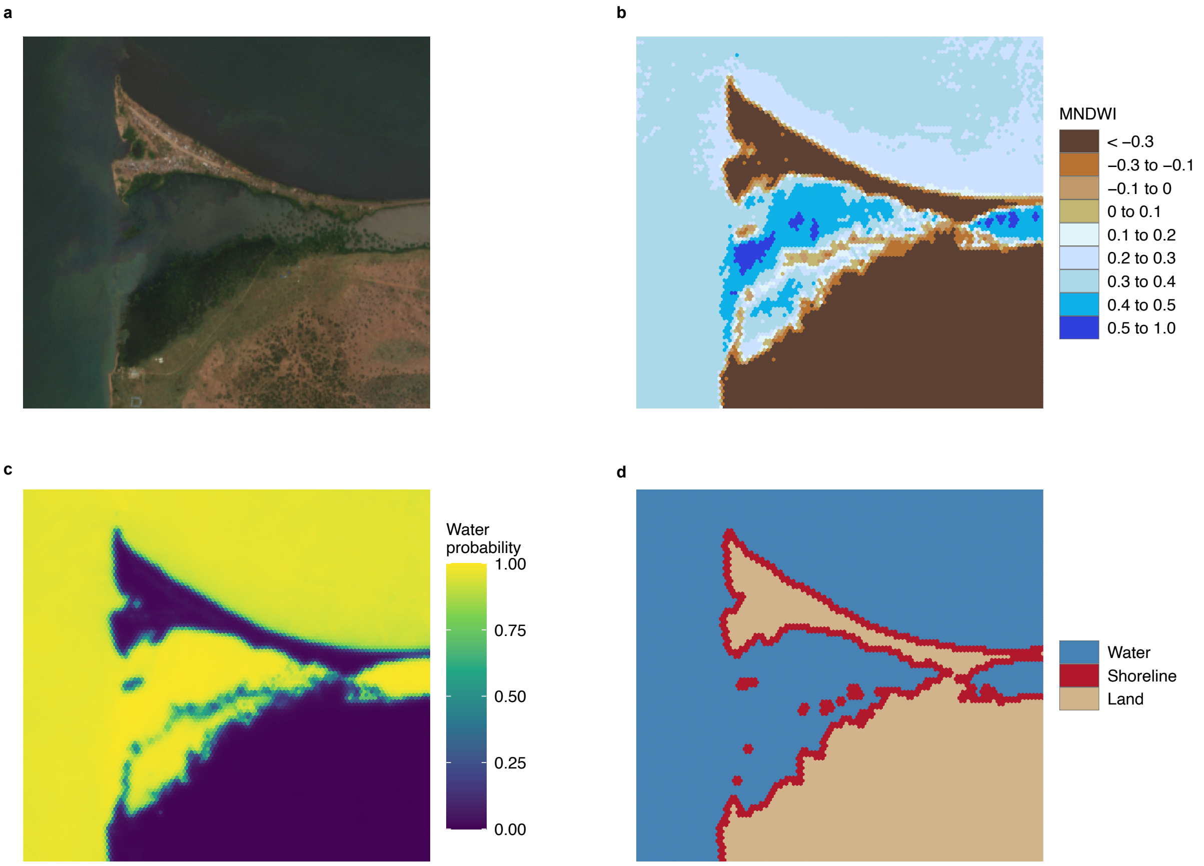


**Fig. S5**: **Satellite-derived shoreline detection.** (**a**) Satellite image of an example area in Buliisa (2023). (**b**) Modified normalised difference water index (MNDWI) from Sentinel-2 on the hexagonal grid. (**c**) Water probability after calibration and smoothing. (**d**) Water and land classification by Otsu's threshold, with shoreline hexagons.

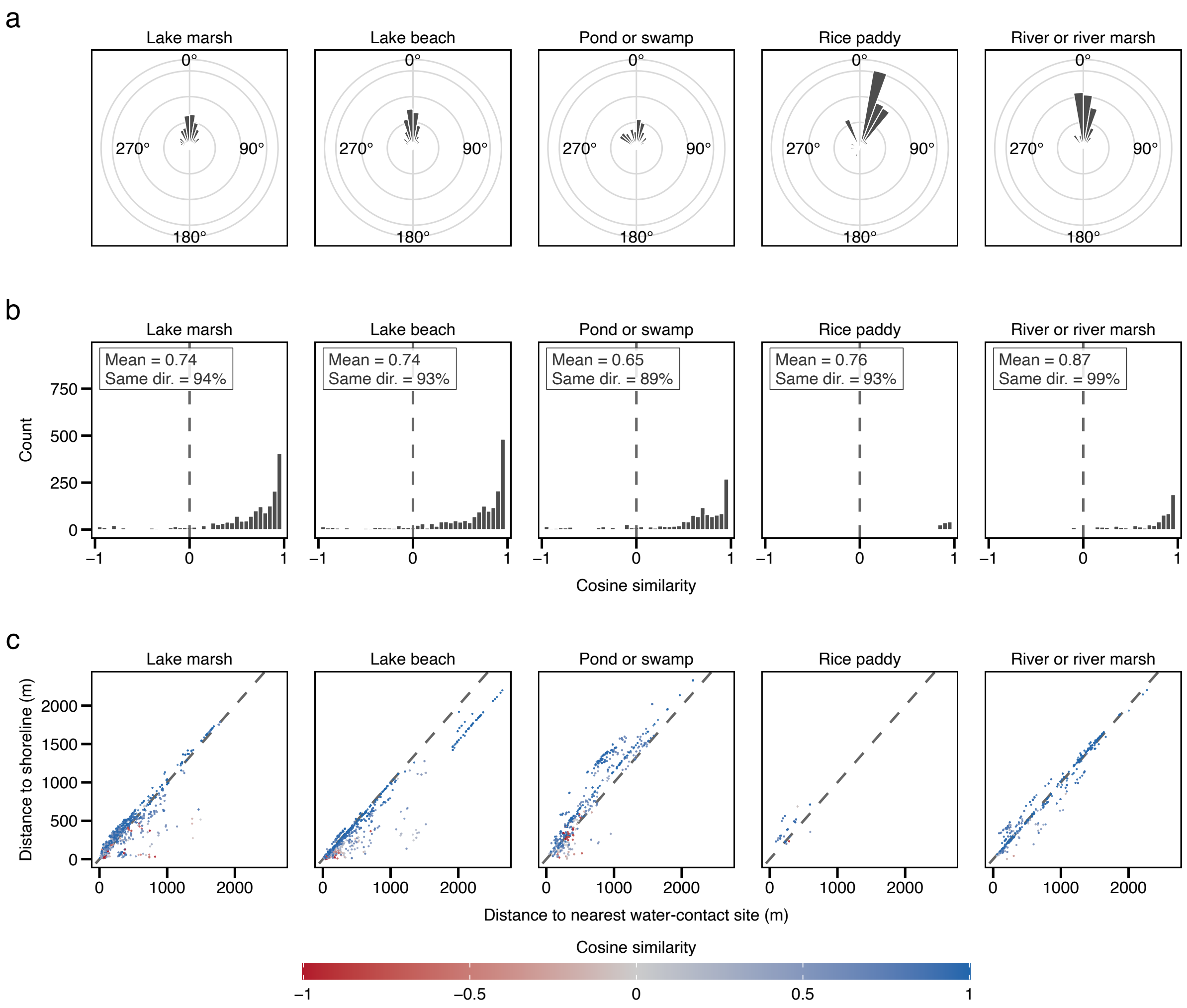


**Fig. S6**: **Validation of the satellite-derived shoreline against field-mapped water-contact sites, by water landform.** (**a**) Direction from each household to the nearest shoreline and to the nearest mapped site. (**b**) Cosine similarity between the two directions. (**c**) Distance to the nearest shoreline against distance to the nearest mapped site, coloured by cosine similarity.

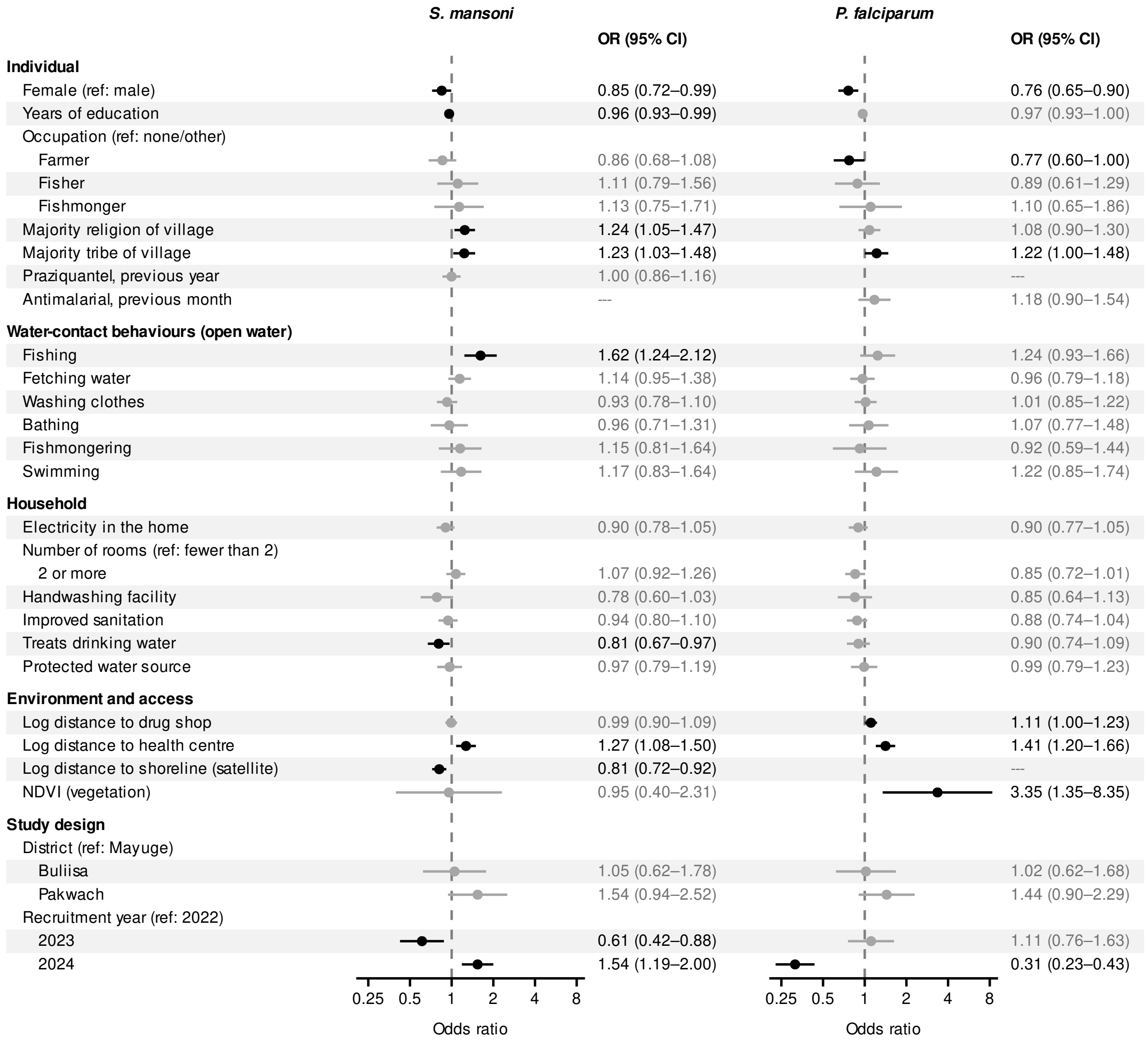


**Fig. S7**: **Adjusted odds ratios from the shoreline model.** Distance to the nearest field-mapped water-contact site replaced by distance to the satellite-derived shoreline and water landform omitted.

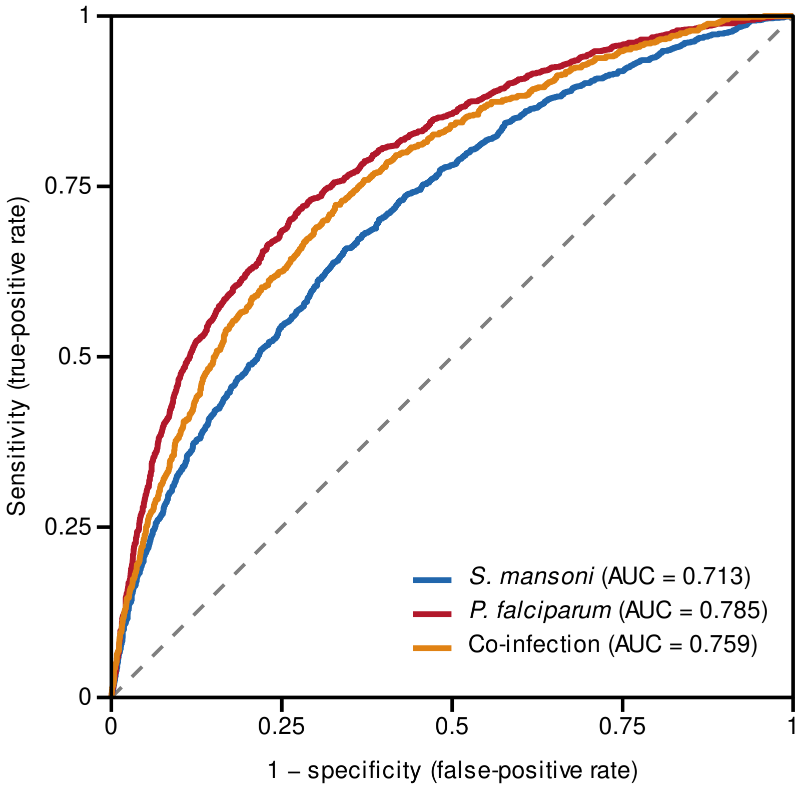


**Fig. S8**: **Receiver operating characteristic curves for the shoreline model under 10-fold cross-validation.** Predictions were pooled across held-out folds. The AUC was 0.713 for *S. mansoni*, 0.785 for *P. falciparum* and 0.759 for co-infection.

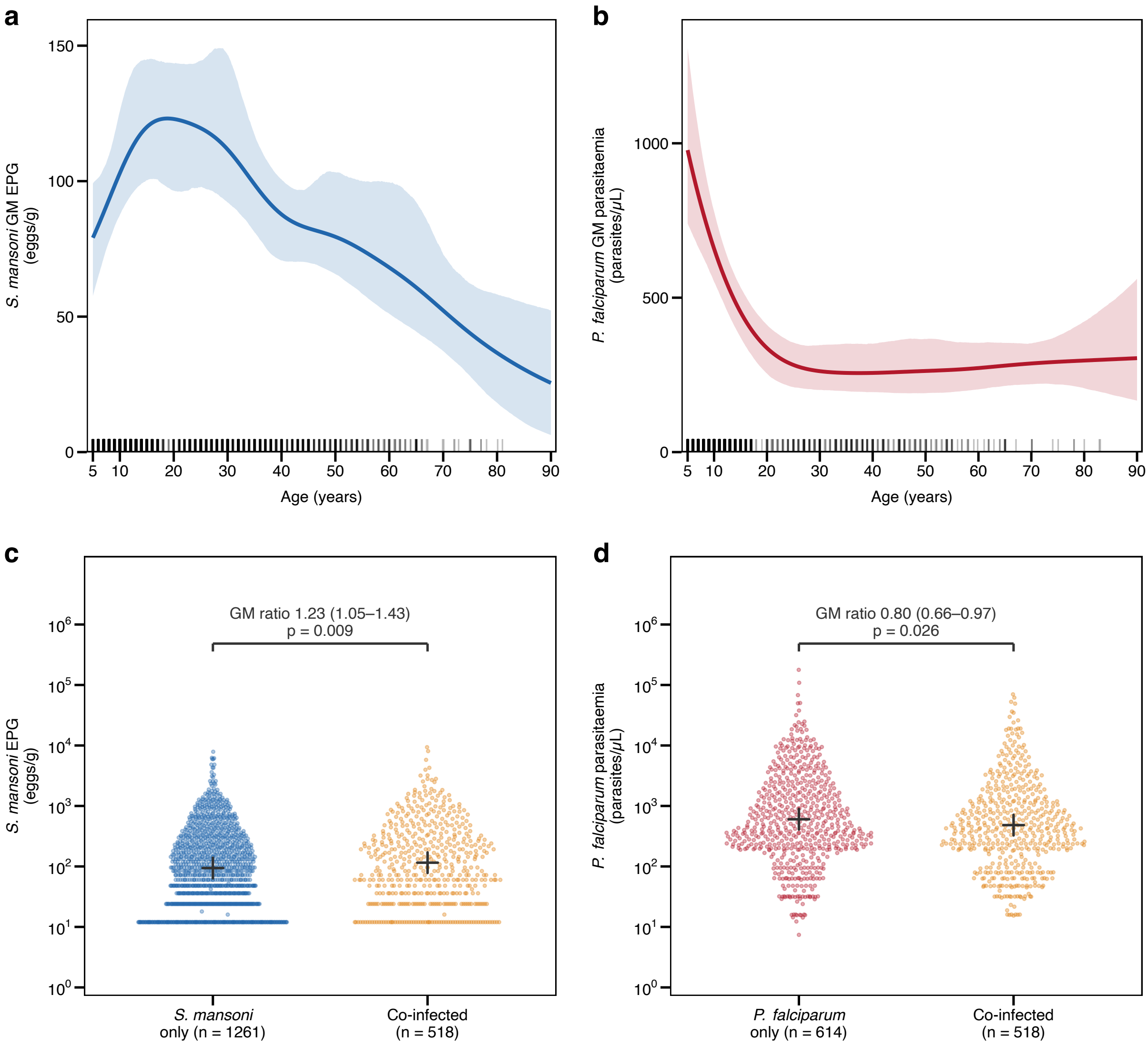


**Fig. S9**: **Infection intensity in mono- and co-infected individuals.** (**a**,**b**) Geometric mean (GM) intensity of *S. mansoni* (EPG) and *P. falciparum* (parasites/$\mu$L) by age, with 95% CIs from a village-level bootstrap. Tick marks show the ages of positive participants. (**c**,**d**) Intensity in mono- and co-infected participants, with *P. falciparum* infection defined by positivity on both RDT and microscopy, so group sizes differ from Table 1. Crosses mark group GMs, and brackets give the GM ratio (co-infected versus mono-infected) with 95% CI and Welch's $t$-test $p$ value.

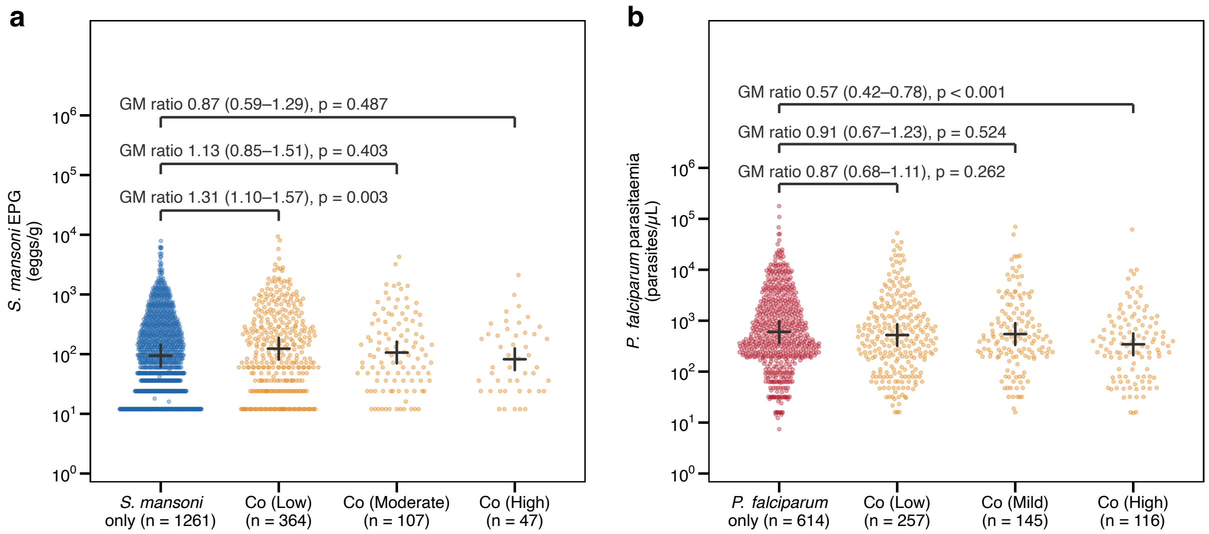


**Fig. S10**: **Infection intensity by load of the co-infecting parasite.** (**a**) *S. mansoni* EPG, with co-infected participants grouped by WHO *P. falciparum* parasitaemia category. (**b**) *P. falciparum* parasitaemia, with co-infected participants grouped by WHO *S. mansoni* intensity category. Each group is compared with mono-infected participants, with markers as in Supplementary Fig. S9.

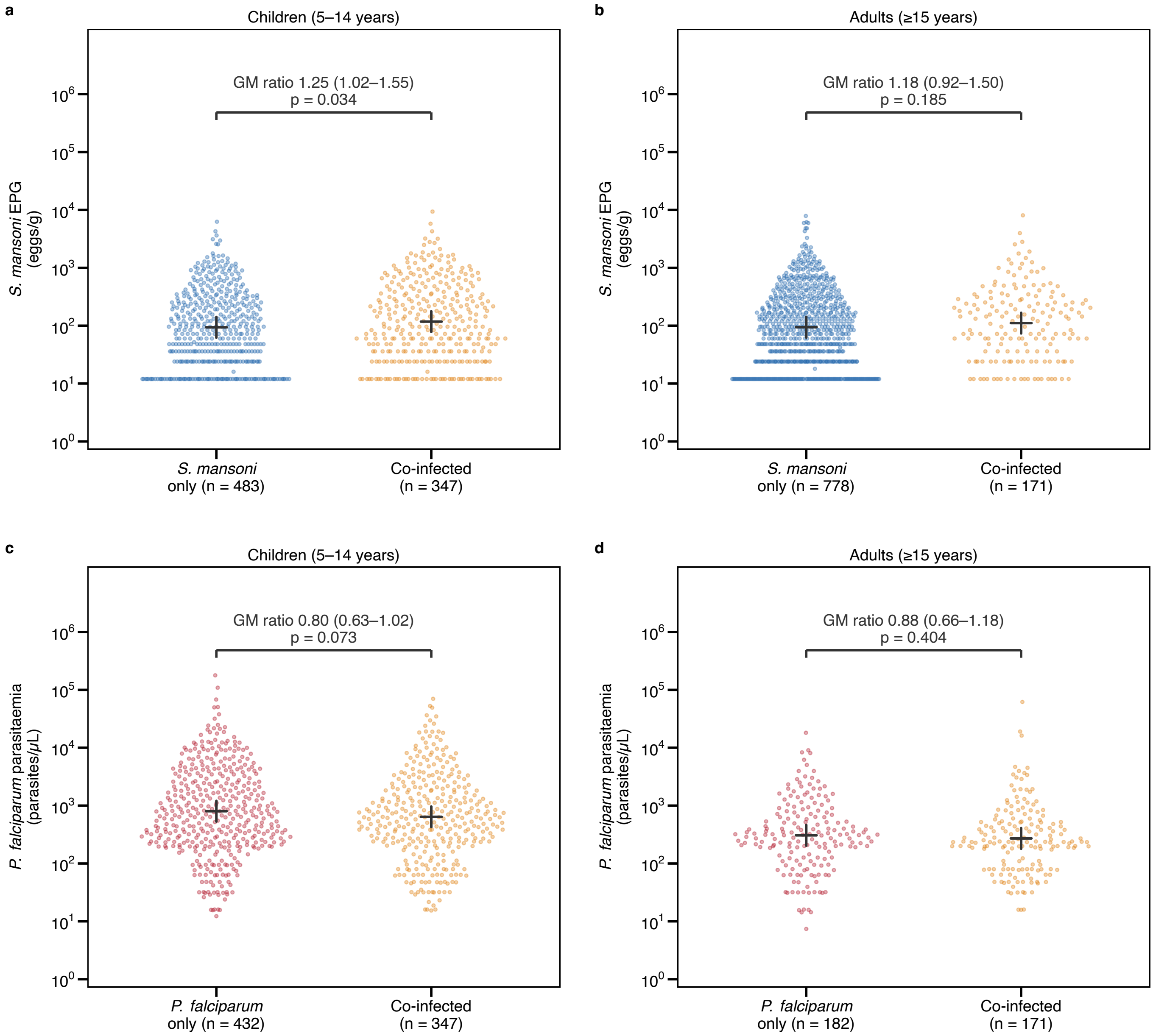


**Fig. S11**: **Infection intensity in mono- and co-infected participants by age group.** *S. mansoni* EPG in (**a**) children aged 5–14 years and (**b**) individuals aged 15 years or older, and *P. falciparum* parasitaemia in (**c**) children and (**d**) individuals aged 15 years or older. Markers as in Supplementary Fig. S9.

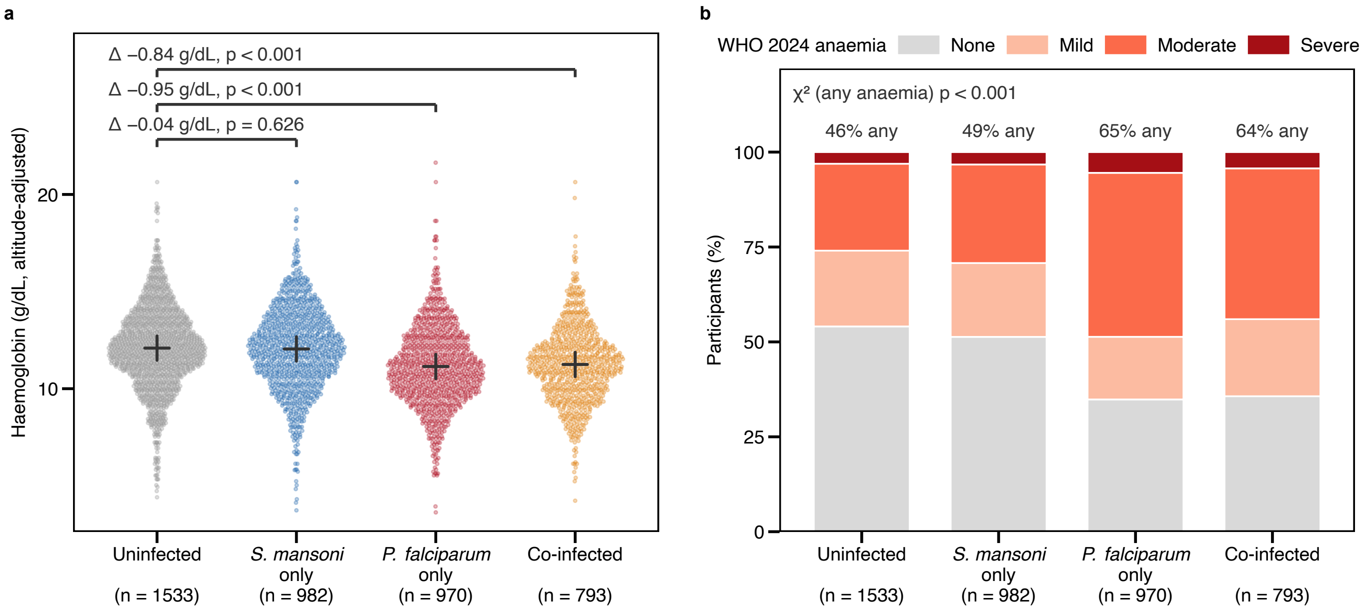


**Fig. S12**: **Haemoglobin and anaemia by infection group** ($n = 4278$). (**a**) Altitude-adjusted haemoglobin. Crosses mark group means, and brackets give the mean difference from uninfected participants (Welch's $t$-test). (**b**) WHO 2024 anaemia categories, with the prevalence of any anaemia compared across groups by $\chi^2$ test. Cutoffs are given in Supplementary Table S10.
